# Longitudinal advanced MRI changes in relapse-free patients with AQP4-IgG+NMOSD

**DOI:** 10.64898/2026.06.18.26355664

**Authors:** Josephine Heine, Darius Mewes, Medha Raman, Patrick Schindler, Klemens Ruprecht, Sven Jarius, Tanja Schmitz-Hübsch, Friedemann Paul, Claudia Chien

## Abstract

Recurrent inflammatory attacks in AQP4-antibody-seropositive neuromyelitis optica spectrum disorder (AQP4-IgG+NMOSD) can lead to devastating disabilities such as visual and motor dysfunction, pain, and cognitive impairment. However, the mechanisms driving the long-term effects of attacks and potential for subsequent recovery are still not well understood after patients enter a relapse-free disease phase.

Here, we leveraged advanced structural and diffusion-weighted imaging analyses in a longitudinal cohort of patients with “stable” AQP4-IgG+NMOSD (retrospectively assessed *≥*12 months without attack, *n*=33, 31/33 female, mean age 49.7 years (SD 14.2)). Brain changes over a median of 4 annual visits (range 2-6) were evaluated using FreeSurfer-based volumetry, regional damage profiles of white matter fibre bundles, cognitive testing (BRB-N), and neuropsychiatric self-reports.

Our analysis revealed four key findings: (1) In the absence of new attacks, pre-existing symptoms persisted and contributed to motor impairment, fatigue, and lower visual function. By contrast, cognitive impairment - selective to higher attention and processing speed - improved over time (PASAT3s, *P_FDR_*=0.007). (2) On a macroscopic brain level, the continued decline of whole brain volumes (*P_FDR_*=0.037) was mainly driven by loss of cortical grey matter (*P_FDR_*=0.013) and linked to poorer motor outcomes (9-hole peg test: *ρₛ*=-0.55, *P_FDR_*=0.021) and higher pain levels (PD-Q: *ρₛ*=-0.51, *P_FDR_*=0.021) at last follow-up. Large-scale age- and sex-stratified reference curves (*Braincharts*) confirmed that cortical atrophy exceeded normal ageing. (3) Thalamic volumes, by contrast, were significantly higher compared to those of healthy participants (*P_FDR_*=0.044) throughout the entire follow-up period and predicted more favourable long-term attention (SDMT: *ρₛ*=0.63, *P_FDR_*=0.003) and spatial memory outcomes (SPART sum score: *ρₛ*=0.62, *P_FDR_*=0.029) as early as at the first MRI. Larger thalamic volumes were mainly seen in a subgroup of younger patients with lower disability burden, fewer comorbidities, and better integrity of thalamus-adjacent white matter tracts. (4) On a microstructural level, tract-specific longitudinal patterns emerged: *decreasing* regional fractional anisotropy (FA) in the optic radiation, thalamo-prefrontal and thalamo-occipital projections was linked to worse cognitive outcomes (e.g., SDMT: *ρₛ*=0.62, *P_FDR_*=0.012), while *increasing* FA, particularly in the corticospinal tract and inferior fronto-occipital fasciculus, predicted more favourable long-term cognitive and visual functions (e.g., NEI VFQ-25: *ρₛ*=0.64, *P_FDR_*=0.011).

Collectively, our data suggest that even in relapse-free AQP4-IgG+NMOSD there is evidence for declining cortical volume, thalamic reserve in some patients, and white matter microstructural damage in distinct regions. Our clinically relevant findings elucidated in the “stable” disease phase highlight longitudinal mechanisms contributing to the long-term prognoses of patients with AQP4-IgG+NMOSD.

## Introduction

Aquaporin-4 immunoglobulin G seropositive neuromyelitis optica spectrum disorder (AQP4-IgG+NMOSD) is a rare autoimmune neuroinflammatory disease.^1,2^ It mainly affects women and has a relapsing disease course with devastating clinical attacks that can result in severe and permanent neurological disability.^3–5^ With better differential diagnostic criteria and understanding of the disease mechanisms, as well as approved attack preventing therapies now available,^6–9^ an unresolved question is whether subtle clinically relevant disease-related neurodegeneration and/or white matter damage occur independent of an acute clinical attack.

Progression independent of relapse activity and silent lesions in the CNS have been shown to be very rare in NMOSD.^10–12^ However, some studies found that patients without an acute AQP4-IgG+NMOSD attack still accrue trans-synaptic and anterograde neurodegeneration^4,13^ and white matter microstructural damage.^14^ These imaging findings could indicate that CNS damage either occurs slowly over time, or that the lack of damage long after a clinical relapse relates to brain and spinal cord plasticity and adaptation to disease-related attacks,^15^ especially since demyelination of the white matter is a secondary event in AQP4-IgG+NMOSD.^15,16^

Initially understood as a disease of the spinal cord and visual system, it is now known that roughly 40% of patients with AQP4-IgG+NMOSD experience cognitive impairment.^17^ Cognitive symptoms can affect several domains, such as higher attention, processing speed, and memory functions.^18,19^ Some studies have linked cognitive impairment in NMOSD to changes in white matter integrity and atrophy of subcortical grey matter structures, such as the thalamus.^20,21^ While thalamic changes are considered a hallmark of cognitive decline in multiple sclerosis,^22,23^ they may be absent in NMOSD^24,25^ or more closely related to visual function^26^ or neuropathic pain.^27^ Atrophy in regional grey matter was shown to mostly involve occipital cortical areas without associated brain lesions.^28^

Our current understanding of how cognition develops over the relapse-free disease course remains incomplete. Repeated cognitive screening in patients with seropositive and seronegative NMOSD revealed that only 10% of the patients had stable scores.^29^ About 30% showed cognitive worsening, while some improved their performance. This opens the possibility that the relapse-free disease phase is not uniform across patients. Instead, there may be an interplay of long-term effects of attacks and subsequent recovery processes. For instance, one cross-sectional study found that lower grey matter brain volume is associated with executive dysfunction in patients with a median relapse-free disease course of 4.9 years.^30^ With this background, we hypothesise that there are subtle, trans-synaptic volumetric and white matter microstructural changes occurring in patients with AQP4-IgG+NMOSD with over a year of relapse-free disease, that are measurable with longitudinal MRI and are related to clinical symptoms and cognitive performance.

To test this hypothesis, we here (I) describe the neuroimaging and clinical profile of patients with NMOSD in the “stable” phase of the disease (attack-free for ≥1 year); (II) examine the evolution of grey matter volume and white matter tract changes in the absence of clinical attacks (particularly in the thalamus and surrounding areas); and (III) determine whether accumulating micro- and macroscopic damage is related to a clinical deterioration in neurological disability and cognitive performance.

## Materials and methods

### Participants and inclusion criteria

Study visits were conducted between 2013-2019 at the NeuroCure Clinical Research Center at Charité–Universitätsmedizin Berlin, Germany (Table 1). Patients were included based on the following criteria: (1) confirmed diagnosis of NMOSD;^1^ (2) seropositive for antibodies against aquaporin-4 in a commercial fixed cell-based assay at any point during the disease course;^31^ (3) ≥18 years of age; (4) attack-free in the year before the MRI; (5) no steroid treatment for at least two months before the MRI, and (6) the availability of multiple MRIs. AQP4-IgG-seronegative patients and patients with significant neurological comorbidities were excluded. Healthy participants were frequency-matched regarding age and sex (Table 2), had no neurological or psychiatric disorders, and received an identical MRI protocol. All participants provided informed written consent. This research was conducted in accordance with the Declaration of Helsinki and approved by the local ethics committee (EA1/041/14 and EA1/163/12).

**Table 1.**
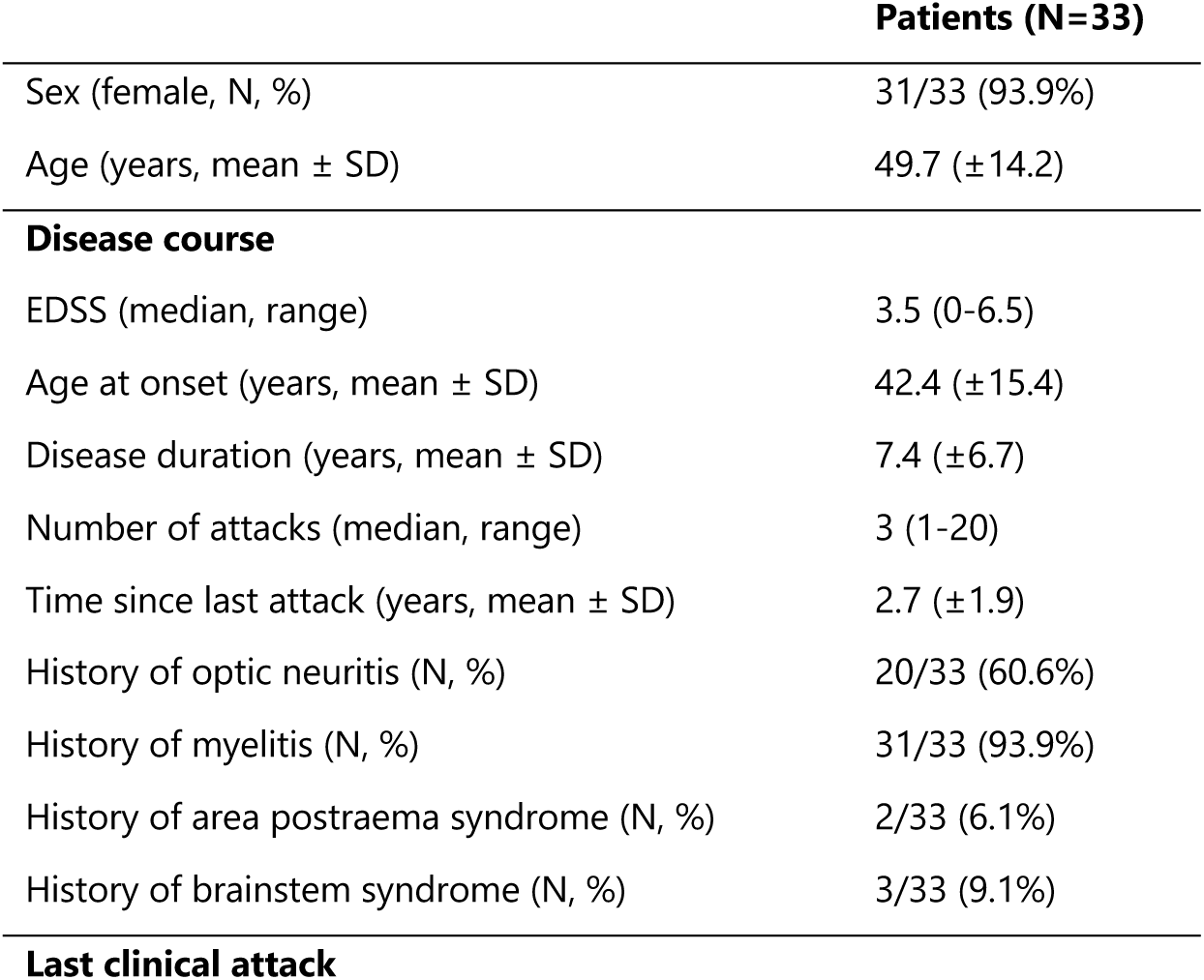
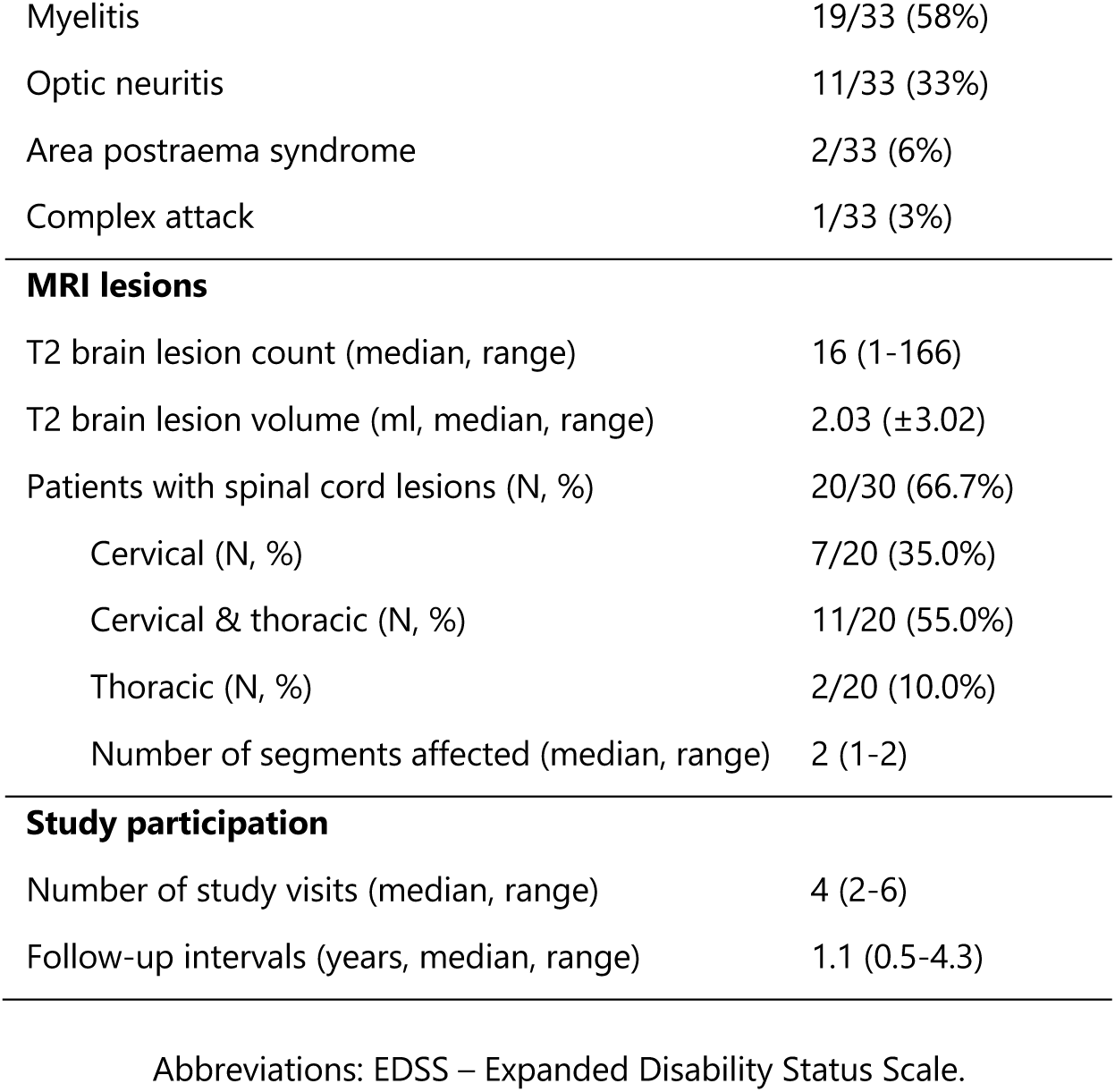
Clinical characteristics.

**Table 2.** Cognitive and functional assessment.

| Assessment (mean ± SD) | Patients (N=33) | Healthy controls (N=27) | First stable visit <sup>a</sup> | Longitudinal development <sup>b</sup> | |
| --- | --- | --- | --- | --- | --- |
| Sex (female, N, %) <sup>c</sup> | 31/33 (93.9%) | 25/27 (92.6%) | $P=1.0$ | - | |
| Age (years) <sup>d</sup> | 49.7 ($\pm 14.2$) | 44.0 ($\pm 12.7$) | $T(57.5)=-1.7$,<br>$P=0.102$ | - | |
| **Clinical symptoms** |  |  |  |  |  |
| MSFC (z-score) | -0.02 ($\pm 0.53$) | 0.39 ($\pm 0.22$) | **$F(1,50)=12.91$,**<br>**$P_{FDR}=0.002$ \*** | $\beta=0.01$ [-0.04,0.05],<br>$P_{FDR}=0.864$ | - |
| 25-foot walk test (sec) | 7.43 ($\pm 5.15$) | 4.35 ($\pm 0.51$) | **$F(1,55)=9.87$,**<br>**$P_{FDR}=0.004$ \*** | $\beta=-0.33$ [-0.60,-0.06],<br>$P_{FDR}=0.089$ | - |
| 9-hole peg test (dominant, sec) | 20.95 ($\pm 3.53$) | 18.50 ($\pm 2.28$) | **$F(1,54)=11.9$,**<br>**$P_{FDR}=0.002$ \*** | $\beta=0.004$ [-0.23,0.23],<br>$P_{FDR}=0.972$ | - |
| 9-hole peg test (non-dominant, sec) | 21.59 ($\pm 4.24$) | 19.03 ($\pm 2.28$) | **$F(1,54)=9.09$,**<br>**$P_{FDR}=0.004$ \*** | $\beta=0.14$ [-0.12,0.40],<br>$P_{FDR}=0.598$ | - |
| Fatigue (FSS) | 3.96 ($\pm 1.90$) | 2.59 ($\pm 1.31$) | **$F(1,55)=9.68$,**<br>**$P_{FDR}=0.004$ \*** | $\beta=0.06$ [-0.05,0.16],<br>$P_{FDR}=0.598$ | - |
| General vision (VFQ25) | 66.00 ($\pm 21.62$) | 91.54 ($\pm 11.56$) | **$F(1,42)=27.7$,**<br>**$P_{FDR}<0.001$ \*** | $\beta=0.40$ [-0.90,1.70],<br>$P_{FDR}=0.812$ | - |
| Current pain (PDQ) | 2.79 ($\pm 2.68$) | - | | | |
| **Cognitive symptoms (BRBN-T)** |  |  |  |  |  |
| PASAT (3 sec) | 43.07 ($\pm 13.49$) | 52.42 ($\pm 5.27$) | **$F(1,50)=10.1$,**<br>**$P_{FDR}=0.020$ \*** | **$\beta=1.21$ [0.51,1.92],**<br>**$P_{FDR}=0.007$ \*** | ↑ |
| SDMT | 49.33 ($\pm 14.48$) | 58.82 ($\pm 7.88$) | $F(1,40)=4.9$,<br>$P_{FDR}=0.076$ | $\beta=0.73$ [-0.11,1.57],<br>$P_{FDR}=0.139$ | - |
| WLG | 26.77 ($\pm 8.41$) | 27.33 ($\pm 6.04$) | $F(1,36)<0.01$,<br>$P_{FDR}=0.851$ | $\beta=0.63$ [-0.03,1.28],<br>$P_{FDR}=0.119$ | - |
| 10/36 SPART sum score (immediate recall) | 19.29 ($\pm 4.53$) | 22.33 ($\pm 3.46$) | $F(1,36)=4.1$,<br>$P_{FDR}=0.079$ | $\beta=0.36$ [-0.01,0.73],<br>$P_{FDR}=0.119$ | - |
| 10/36 SPART-D (delayed recall) | 7.71 ($\pm 2.07$) | 9.22 ($\pm 1.09$) | $F(1,36)=5.0$,<br>$P_{FDR}=0.076$ | $\beta=-0.01$ [-0.18,0.15],<br>$P_{FDR}=0.884$ | - |
| SRT-LTS (immediate recall) | 52.68 ($\pm 13.08$) | 60.56 ($\pm 7.86$) | $F(1,36)=4.6$,<br>$P_{FDR}=0.076$ | $\beta=0.75$ [-0.21,1.74],<br>$P_{FDR}=0.151$ | - |
| SRT-CLTR (immediate recall) | 46.55 ($\pm 16.69$) | 54.56 ($\pm 14.03$) | $F(1,36)=2.5$,<br>$P_{FDR}=0.142$ | $\beta=1.11 [-0.04,2.27]$,<br>$P_{FDR}=0.119$ | – |
| SRT-D (delayed recall) | 10.45 ($\pm 1.93$) | 11.33 ($\pm 1.12$) | $F(1,36)=2.6$,<br>$P_{FDR}=0.142$ | $\beta=-0.07 [-0.20,0.06]$,<br>$P_{FDR}=0.322$ | – |

<sup>a</sup> ANCOVA adjusted for age and sex. <sup>b</sup> Linear mixed effects models for time since last attack, adjusted for age and sex. <sup>c</sup> Fisher's exact test. <sup>d</sup> Student's t-test. Abbreviations: BRBN-T – Brief Repeatable Battery of Neuropsychological Tests, CLTR – consistent long-term retrieval, FSS – Fatigue Severity Scale, LTS – long-term storage, MSFC – Multiple Sclerosis Functional Composite, PASAT – Paced Auditory Serial Addition Test, PDQ – PainDETECT Questionnaire, SD – standard deviation, SDMT – Symbol Digit Modalities Test, SPART – Spatial Recall Test, SRT – Selective Reminding Test, VFQ25 – Visual Function Questionnaire 25, WLG – Word List Generation.

### Clinical and cognitive data

Functional outcomes were evaluated as standardised z-scores using the Multiple Sclerosis Functional Composite Score (MSFC).^32,33^ Additionally, we assessed fatigue using the Fatigue Severity Scale (FSS),^34^ current neuropathic pain (painDETECT Questionnaire (PD-Q), “How would you assess your pain now, at this moment?”),^35^ and self-reported visual function (National Eye Institute Visual Function Questionnaire (NEI VFQ-25), “At the present time, would you say your eyesight using both eyes (with glasses or contact lenses, if you wear them) is excellent, good, fair, poor, or very poor or are you completely blind?”).^36^ Cognition was assessed using the Brief Repeatable Battery of Neuropsychological Tests (BRBN-T),^37^ including higher attention and processing speed (Paced Auditory Serial Addition Test, PASAT;^38^ Symbol Digit Modalities Test, SDMT),^39^ semantic fluency (Word List Generation, WLG),^37^ as well as visuospatial (10/36 Spatial Recall Test, SPART)^40^ and verbal learning and recall (Selective Reminding Test, SRT).^41^

### MRI data acquisition

All MRI images were acquired on the same 3T Siemens MAGNETOM Tim Trio machine (Siemens, Erlangen) equipped with a 32-channel head coil at the Berlin Center of Advanced Neuroimaging (BCAN). The imaging protocol included a 3D T1-weighted MPRAGE (1mm³, Supplementary Methods), a 3D FLAIR (1mm³), a single-shot EPI diffusion-weighted imaging sequence (DWI), as well as a 2D-sagittal T2-weighted SC sequence at the cervical, thoracic, and lumbar level of the spinal cord. All follow-up assessments were conducted using an identical imaging protocol and test battery.

### MRI data analysis

#### Brain volumetry

Volumetric brain parcellations were performed using T1-weighted MPRAGEs and the longitudinal stream in FreeSurfer 6.0.^42^ This automated approach reduces random variation by creating an unbiased template for each participant. Based on the within-subject template, FreeSurfer performs brain extraction, Talairach transformation, atlas registration, and parcellation, allowing to extract volumes with increased reliability.^43^ We collected whole brain volumes, including total grey matter, white matter, and cortex, as well as volumetric estimates of subcortical grey matter structures, the choroid plexus, and cerebellum. Moreover, we performed a brainstem subfield segmentation into midbrain, pons, superior cerebellar peduncle, and medulla oblongata.^44^ Individual parcellations underwent visual quality control. If necessary, manual corrections of automated parcellations were performed to ensure the validity of outputs.

#### Normative volumetric trajectories using Braincharts

In addition to the regional segmentations described above, we obtained normative centiles for patients with AQP4-IgG+NMOSD based on Braincharts reference charts (https://www.brainchart.io), the largest normative dataset for volumetric phenotypes of the human brain to date.^45^ Here, we benchmarked total cortical volume, total white matter volume, and total subcortical grey matter volume against normative data across 101 studies based on FreeSurfer 6.0. For the age range of this study, the normative values were aggregated from *n*=95,536 healthy participants out of a total of *n*=101,457 individuals.

#### Brain T2-weighted lesion segmentation

Manual T2-hyperintense brain lesions were segmented by MRI technicians with ≥15 years of experience from baseline cerebral FLAIR MRIs in Montreal Neurological Institute 152 (MNI-152) space, creating binary lesion masks using ITK-SNAP.^46^ The lesion masks were further subsegmented with different labels indicating regions relevant for NMOSD, as described in Mewes et al. (in preparation). Briefly, the lesion masks were subsegmented into eight different NMOSD-relevant categories fitting into the periventricular, infratentorial, juxtacortical, and subcortical regions (see Supplementary Methods).^1^

### Diffusion-weighted imaging analysis

#### White matter fibre-bundle extraction

DWI from each patient session were pre-processed as described in Mewes et al. (in preparation). Briefly, DWI removal of Gibb’s ringing, distortion correction, brain extraction of the B0 volume, and fitting of the diffusion tensor model were performed with MRtrix3 (v.3.0.3)^47^ and FSL libraries.^48^ White matter tract templates for each patient were generated and non-linearly transformed to IIT-Atlas space using DTI-TK.^49^

#### Regional fibre-bundle fractional anisotropy profiles

White matter tracts of high interest in “stable” AQP4-IgG+NMOSD were selected for fibre-bundle analysis.^50^ Fractional anisotropy (FA) was calculated using dtifit (https://fsl.fmrib.ox.ac.uk/fsl/docs/diffusion/dtifit.html). Bundle-specific tractograms^51^ were generated based on TractSeg atlas definitions, resampled to equidistant slices, and divided into two or three regions of interest. The number of slices dependended on the distances between median endpoint coordinates of each tractogram, which were divided by the voxel size of the DWI sequence and rounded to the nearest ten. FA values were sampled per slice using *tcksample* and aggregated into mean FAs for each region.

#### Longitudinal fibre-bundle regional damage calculation

Patient group mean FA extracted from each region of white matter fibre-bundles were compared between each consecutively available MRI visit with the first available MRI with coinciding clinical visit. On the individual patient-level, calculation of damage in a fibre-bundle was carried out by a histogram method by Mewes et al. (in preparation), an extension of that proposed by Stamile et al.^52^ It should be noted that the “Visit” label was based on the clinical visit, thus, not all timepoints contained the same number of patient FA values per region due to missing DWI scans at certain clinical visits. One patient only had one DWI scan, thus was excluded from further longitudinal white matter analysis.

Briefly, distributions of streamline FA values for each patient, bundle, and slice, at baseline and follow-up were modelled using Gaussian Mixture Models. A per-slice threshold indicating relevant longitudinal FA decrease (as in demyelination) was calculated using an optimization procedure. Subsequently, slice damage was computed as the proportion of streamlines below the FA threshold. To reduce false-positive findings, the smallest real difference (SRD;^53^ Equation 1) was derived from damage values in the lowest slice of the corticospinal tract, comparing the first follow-up DWI scan to baseline across patients in the relapse-free AQP4-IgG+NMOSD cohort. This region was chosen because it is particularly prone to MRI signal drop-out. Then, for each anatomical bundle region, slice-wise damage values were averaged to obtain regional damage estimates, with slices below the SRD contributing zero to the regional mean:

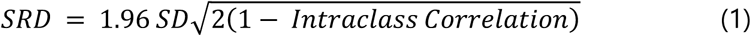

where SD is the standard deviation. The thresholded mean damage in a fibre-bundle region per patient was determined between each consecutive DWI session (Equation 2):

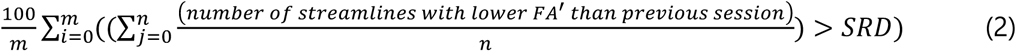

where m is the number of slices per region, n is the number of streamlines per slice in a tractogram, and FA’ is the FA decrease identified by the optimised GMM (see Supplementary Methods).

#### Spinal cord imaging

From the 2D spinal cord MRIs, T2-weighted lesions were identified and counted, including vertebral length. We estimated the mean upper cervical cord area (MUCCA) for the first available 3D MPRAGE scan as described previously.^54,55^ Briefly, JIM 7.0 (http://www.xinapse.com) was used to average over five consecutive slices with the C2-C3 intervertebral space as a landmark.

### Statistical analysis

Statistical analyses were performed in R4.5.0. (R Core Team, 2025, https://www.r-project.org/). For cross-sectional analyses, we compared volumetric markers of the first “stable” MRI between patients and matched healthy participants using ANCOVAs adjusted for sex, age, and estimated intracranial volume (eTIV). Effect sizes are reported as partial *η²* (Cohen, 1988).^56^

Longitudinal volumetric trajectories were analysed using linear mixed effects models (*lmer*).^57^ Here, we modelled time since last attack as predictor, the MRI variable as outcome, and sex, age, and intracranial volume as fixed effects. Additionally, we adjusted for different starting points between individuals using random intercepts. For group white matter fibre-bundle comparisons, mean differences were calculated using 5000 bootstrapped samples and effect sizes (Cohen’s *d*) were used as an indicator of true population distributions in DWI-extracted FA (*dabestr*). Cross-sectional associations with clinical variables were analysed using multiple linear regression models (predictors: age at onset, time since onset, time since last attack, number of attacks, EDSS, current pain level (PD-Q), PASAT3s score). To analyse change over time, we calculated delta scores, e.g., Δ_brain volume_ = volume_(last visit)_ – volume_(first visit)_. Lastly, outcomes were predicted using Spearman correlations of delta scores with clinical scales at last study visit, e.g., Δ_brain volume_ ∼ EDSS_(last visit)_.

Descriptive statistics are presented as mean (±SD), median (range), and proportions (95% confidence intervals). Group comparisons were adjusted for multiple testing using the Benjamini-Hochberg correction based on the *p.adjust* package in R. Where applicable, two-tailed tests were used. An alpha level of <0.05 was considered statistically significant. Figures were created using the R packages *ggplot2, dabestr, ggpubr,* and *smplot*.^58,59^

## Results

### Sample selection & clinical characteristics

We identified *n*=33 patients fulfilling the inclusion criteria after screening *n*=82 patients with *n*=244 sessions (*n*=32 with longitudinal DWI). The last clinical attack occurred on average 2.7 years (*SD*=1.9 years, range 1.0-9.8 years) before the first MRI. Across the follow-up, almost all patients received concurrent treatments (30/33, 91%) at some point, either alone or in combination with treatment for pain (13/33, 39%), muscle spasms (6/33, 18%), or anxiety and depression (7/33, 21%). Half of the patients presented with comorbidities (not meeting exclusion criteria, 18/33, 55%). Most study participants self-identified as White/European (*n*=31). One patient identified as Black/African, and one patient reported Arab descent. Further details on demographic details, treatments, comorbidities, and study exclusion are provided in Supplementary Table 1.

### Clinical and cognitive outcomes

At first evaluation during the “stable” period (median 1.8 years after last attack), patients experienced significantly reduced walking speed and mobility (25-foot walk test, *P_FDR_*=0.004, *η²ₚ*=0.15) and reduced fine motor and upper extremity function compared to healthy participants (9-hole peg test, dominant hand: *P_FDR_*=0.002, *η²ₚ*=0.18, non-dominant hand: *P_FDR_*=0.004, *η²ₚ*=0.14, Table 2). Moreover, they reported higher levels of fatigue (FSS, *P_FDR_*=0.004, *η²ₚ*=0.15) and lower quality of life related to visual function (VFQ25, *P_FDR_*<0.001, *□²ₚ*=0.40, all large effects). None of these functional outcomes improved or deteriorated over the follow-up time.

Processing speed and higher attention were affected, with lower scores on the PASAT compared to healthy participants (*P_FDR_*=0.017, *η²ₚ*=0.18, large effect). Over a median of 4 years with annual testing, the PASAT performance improved significantly (*P_FDR_*=0.007). Moderate effects were also seen for impairments on the SDMT (*η²ₚ*=0.11), as well as for visual (SPARTsum: *η²ₚ*=0.10, delayed recall: *η²ₚ*=0.12) and verbal memory performance (SRT-LTS: *η²ₚ*=0.11, delayed recall: *η²ₚ*=0.07), albeit not statistically significant after correction for multiple testing. No further longitudinal changes were noted.

### Volumes

#### Normative volumetric analysis (Braincharts)

Individual centile scores for global brain volumes were estimated based on the age- and sex-stratified normative reference curves (mean age 40.03 years). Here, cortical grey matter showed a significant longitudinal decrease in patients with relapse-free AQP4-IgG+NMOSD (*ß*=-1.00 [-1.81,-0.19], *P_FDR_*=0.049, Fig. 1). Lower initial grey matter centile scores were linked to worse cognitive outcomes at last follow-up, however, this association did not remain significant after correction for multiple testing (PASAT3s: *ρₛ*=0.44, *P_FDR_*=0.089, SPART delayed recall: *ρₛ*=0.56, *P_FDR_*=0.089). In contrast, the cerebral white matter (*ß*=-0.27 [-0.97,0.43], *P_FDR_*=0.635) and total subcortical grey matter volumes (*ß*=0.21 [-0.66,1.08], *P_FDR_*=0.635) did not show any accelerated volume decline outside of normal ageing.

**Figure 1.**
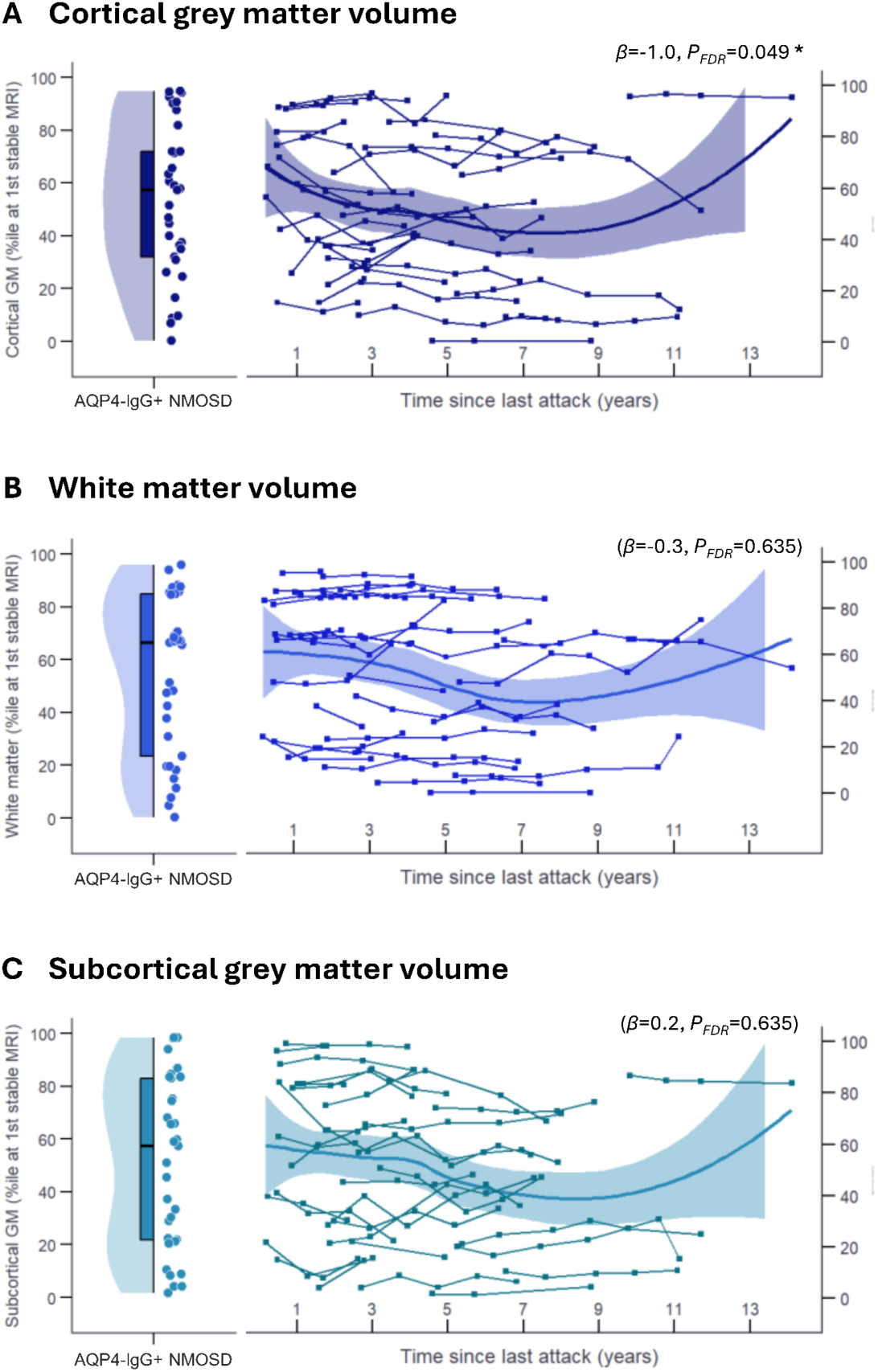
Normative centiles for patients with stable AQP4-IgG+NMOSD based on age- and sex-stratified reference charts (*Braincharts*, A-C). Abbreviation: GM – grey matter.

#### Whole brain volumes

At a median of 1.8 years since the last attack, patients with AQP4-IgG+NMOSD showed distinct global volume reductions compared to healthy participants (Table 3) for the whole brain (*P_FDR_*=0.001, *η²ₚ*=0.26), white matter (*P_FDR_*=0.002, *η²ₚ*=0.23), and grey matter (*P_FDR_*=0.022, *η²ₚ*=0.13), all with medium to large effect sizes (Fig. 2). In particular, cortical grey matter volume was lower in patients (*P_FDR_*=0.028, *η²ₚ*=0.12) and, like total grey matter volume, significantly linked to older age of onset (*β*=-1,758, *SE*=536, *P_FDR_*=0.010) and a higher number of attacks (*β*=-7,730, *SE*=2,218, *P_FDR_*=0.009, *R²_adj_*=0.41). Moreover, lower whole brain volumes were significantly associated with lower cognitive scores (PASAT3s: *β*=2,768, *SE*=1,079, *P_FDR_*=0.048) and a higher number of attacks (*β*=-14,722, *SE*=5,198, *P_FDR_*=0.040, *R²_adj_*=0.31). No difference was found for total subcortical grey matter.

**Figure 2.**
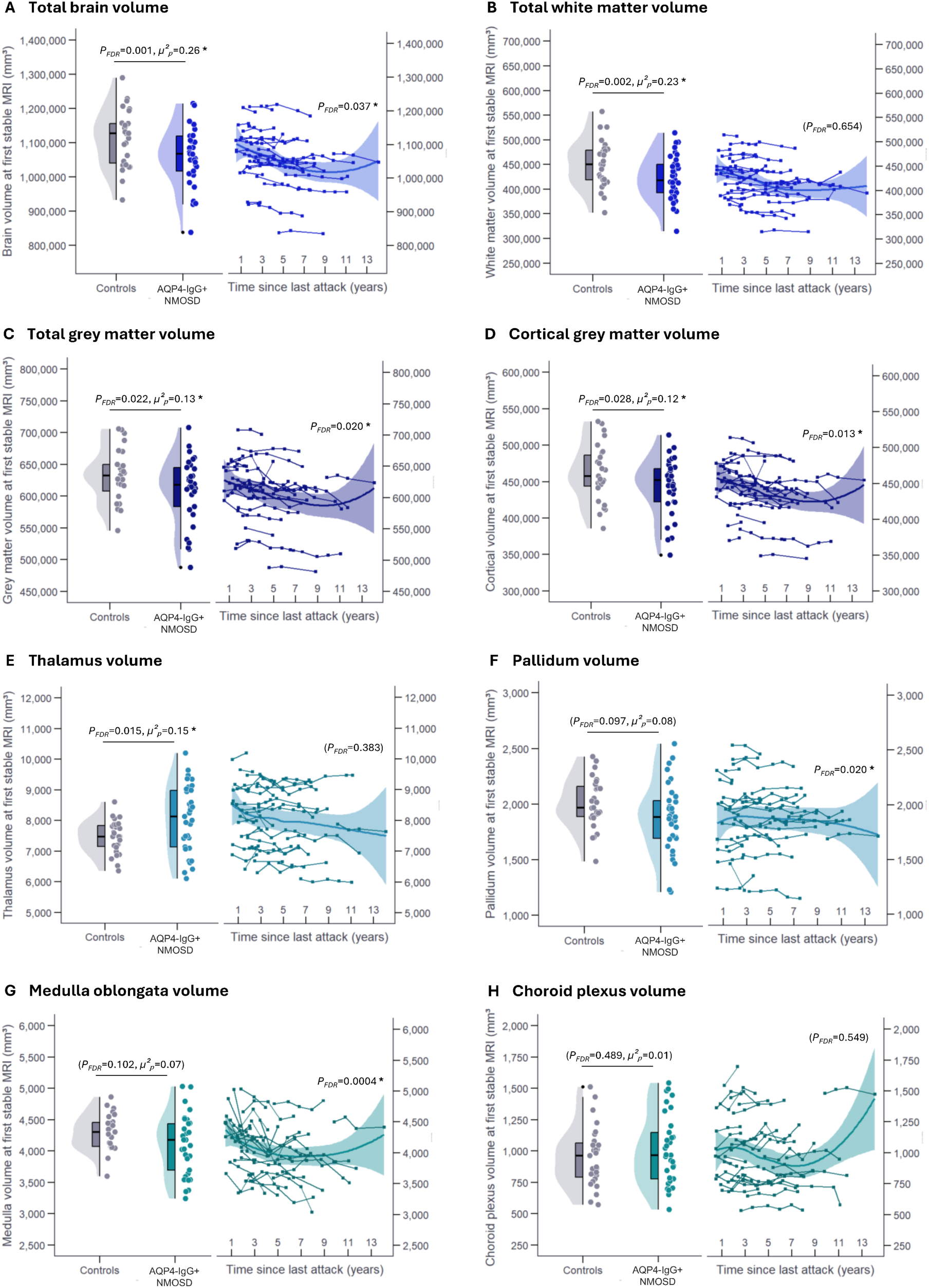
Global and regional volumetric outcomes in patients with stable AQP4-IgG+NMOSD (A-H).

**Table 3.**
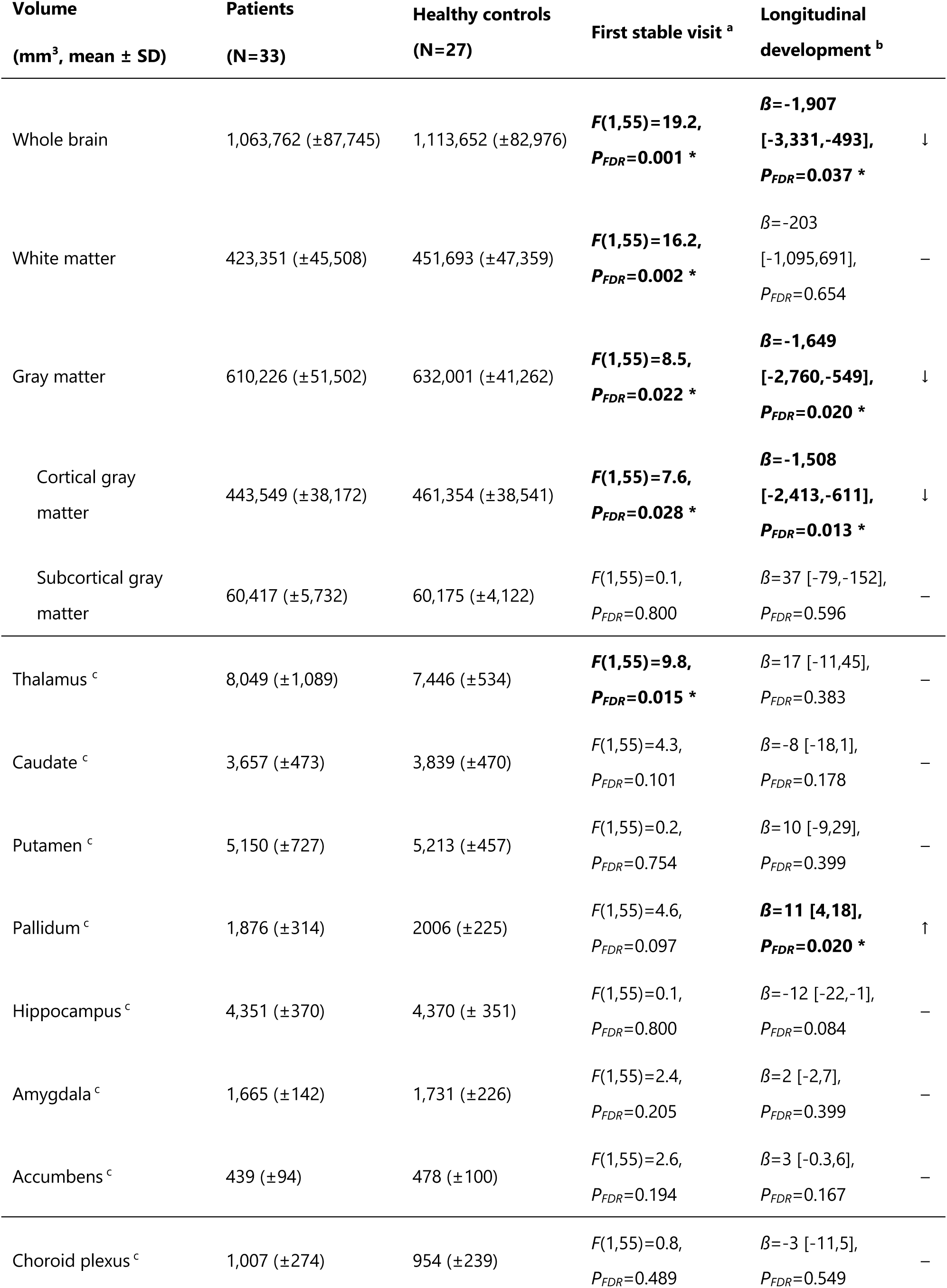
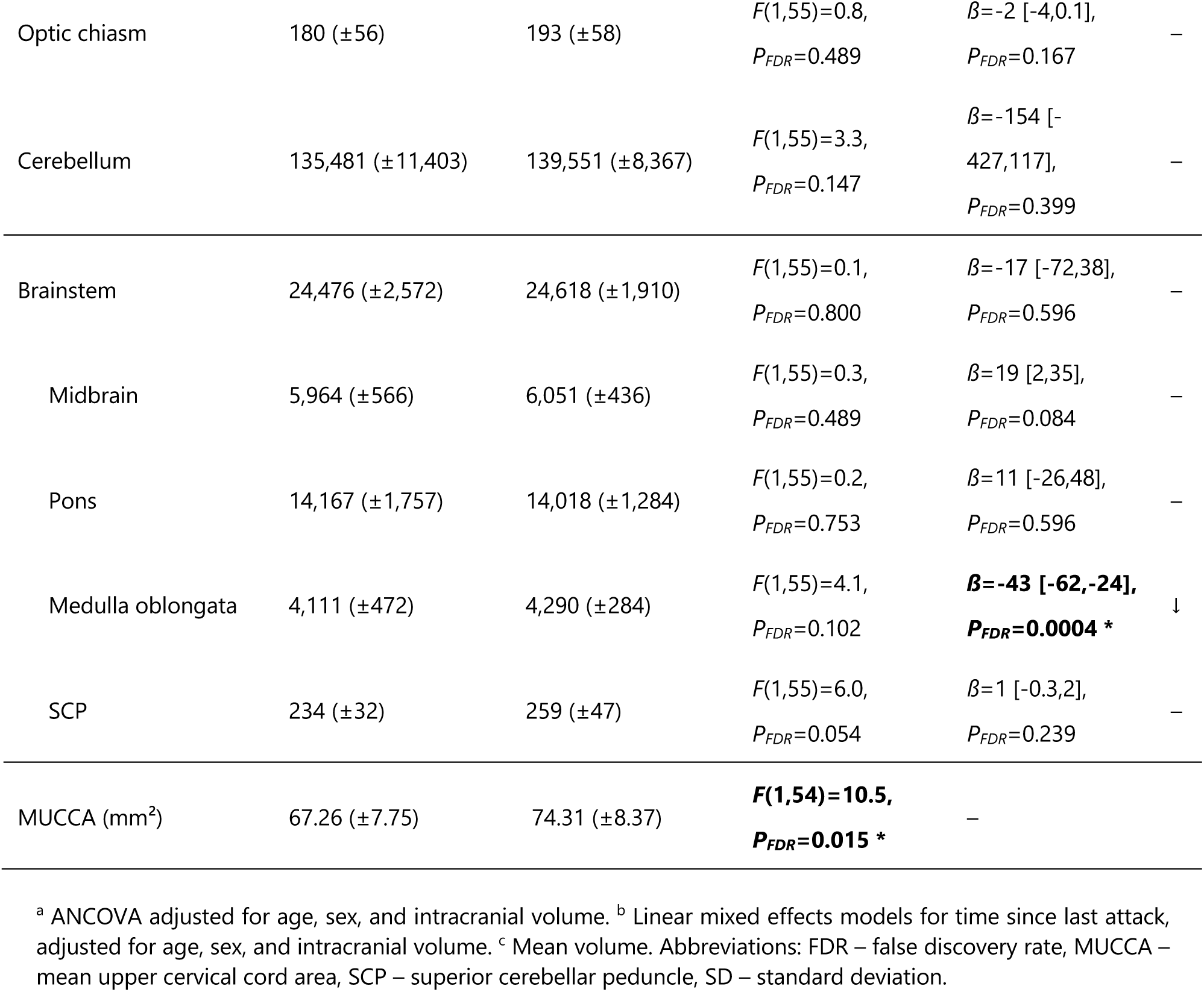
Brain volume estimates.

Over time, whole brain (*P_FDR_*=0.037), total grey matter (*P_FDR_*=0.020), and cortical grey matter volumes (*P_FDR_*=0.013) continued to decrease. White matter volume remained stable over time. More extensive longitudinal brain volume loss was significantly associated with worse motor outcomes (9-hole peg test: *ρₛ*=-0.55, *P_FDR_*=0.021) and higher pain levels at last follow-up (PD-Q: *ρₛ*=-0.51, *P_FDR_*=0.021). Although not significant after correction for multiple comparisons, greater volume decline also tended to correlate with worse cognitive outcomes on tests of attention (PASAT3s: *ρₛ*=0.49, *P_FDR_*=0.062, SDMT: *ρₛ*=0.45, *P_FDR_*=0.062).

#### Thalamus volume

Initially, patients with relapse-free AQP4-IgG+NMOSD showed increased thalamic volumes (*P_FDR_*=0.015, *µ²ₚ*=0.15, large effect) compared to healthy participants. These were significantly associated with a younger age at onset (*β*=-41.6, *SE*=15.1, *P_FDR_*=0.044) and fewer attacks (*β*=-162.8, *SE*=62.5, *P_FDR_*=0.044, *R²_adj_*=0.38). Larger thalamic volumes remained stable over time (Table 3). As early as at the first stable MRI, thalamic volumes predicted better subsequent cognitive outcomes at last follow-up, including higher attention (SDMT: *ρₛ*=0.63, *P_FDR_*=0.003) and spatial memory (SPARTsum: *ρₛ*=0.62, *P_FDR_*=0.029).

#### Subcortical volumes, brainstem volumes, and spinal cord cross-sectional area

No differences were observed for other subcortical structures (Table 3). The pallidum, despite no initial difference, showed a longitudinal volume *increase* in “stable” patients (*P_FDR_*=0.020, Fig. 2). In contrast, volume estimates of the medulla oblongata *decreased* significantly over time (*P_FDR_*=0.0004). While we observed a moderate effect for lower cross-sectional volumes of the medulla oblongata (*η²ₚ*=0.07) and superior cerebellar peduncle (*η²ₚ* =0.10) at first stable MRI, these brainstem subfield volumes did not survive correction for multiple comparisons (*P_FDR_*=0.102 and *P_FDR_*=0.054, respectively). Moreover, the mean upper cervical cord area (MUCCA) was significantly smaller in patients compared to healthy controls (*P_FDR_*=0.015, *η²ₚ*=0.16, large effect). No associations with clinical scores were observed after correction for multiple comparisons (Supplementary Fig. 1).

### Brain lesions

Not surprisingly, the most common lesions in patients at the first available MRI were non-specific white matter lesions, with a median count of four (95% *CI*: 0.69 to 0.95) and mean volume of 0.469 (±0.768) mL. The second most observed lesion location was by the lateral ventricles with a median of three lesions (95% *CI:* 0.51 to 0.83) and mean volume of 1.244 (±2.094) mL. Apart from individual juxtacortical lesions (median count: 1, 95% *CI:* 0.38 to 0.72, mean volume: 0.127 (±0.245) mL), little to no lesions were found elsewhere in the brain (see Supplementary Table 2). Longitudinally, both lesion count (*ß*=-0.37 [-1.35,0.62], *P_FDR_*=0.490) and volume (*ß*=-0.03 [-0.11,0.01], *P_FDR_*=0.490) remained stable. A low lesion volume early in the “stable” phase was associated with better visual memory performance at last follow-up (*ρₛ*=-0.73, *P_FDR_*=0.007). No associations between lesions and motor outcomes or disease course variables were found.

### White matter microstructural analysis

#### Regional fibre-bundle fractional anisotropy

At the first available DWI, we observed relatively low regional FA in several fibre bundles in patients with AQP4-IgG+NMOSD, although within the normal range for the mean age of our cohort (Fig. 3A).^60^ The following tracts showed FA values lower than 0.5: the left corticospinal tract (CST, cortical region, mean FA (±SD): 0.40 (±0.03)), the right optic radiation (OR, 0.39 (±0.04)), as well as the left (anterior region: 0.42 (±0.02), posterior region: 0.41 (±0.05)) and right (anterior region: 0.41 (±0.02)) inferior fronto-occipital fasciculus (IFOF). Similarly, the thalamic projections fibres showed a tendency to have lower FA, including the left (anterior region: 0.37 (±0.03), posterior region: 0.38 (±0.02)) and right (anterior region: 0.34 (±0.04), posterior region: 0.27 (±0.02)) thalamic projections to the prefrontal cortex (TPFC), along with the right thalamic projection to the occipital lobe (posterior region, TPOL, 0.41 (±0.04)). Lower FA in the left posterior TPFC was significantly associated with higher disability (*β*=-0.010, *SE*= 0.003, *P_FDR_*=0.026). Moreover, lower FA in the right posterior TPFC was associated with worse motor function (9-hole peg test: *ρₛ*=-0.71, *P_FDR_*=0.0003), as well as worse performance on tests on higher attention (SDMT: *ρₛ*=0.58, *P_FDR_*=0.009), visuospatial (SPARTsum: *ρₛ*=0.44, *P_FDR_*=0.042) and verbal memory (SRTsum: *ρₛ*=0.47, *P_FDR_*=0.042, Supplementary Fig. 2).

**Figure 3.**
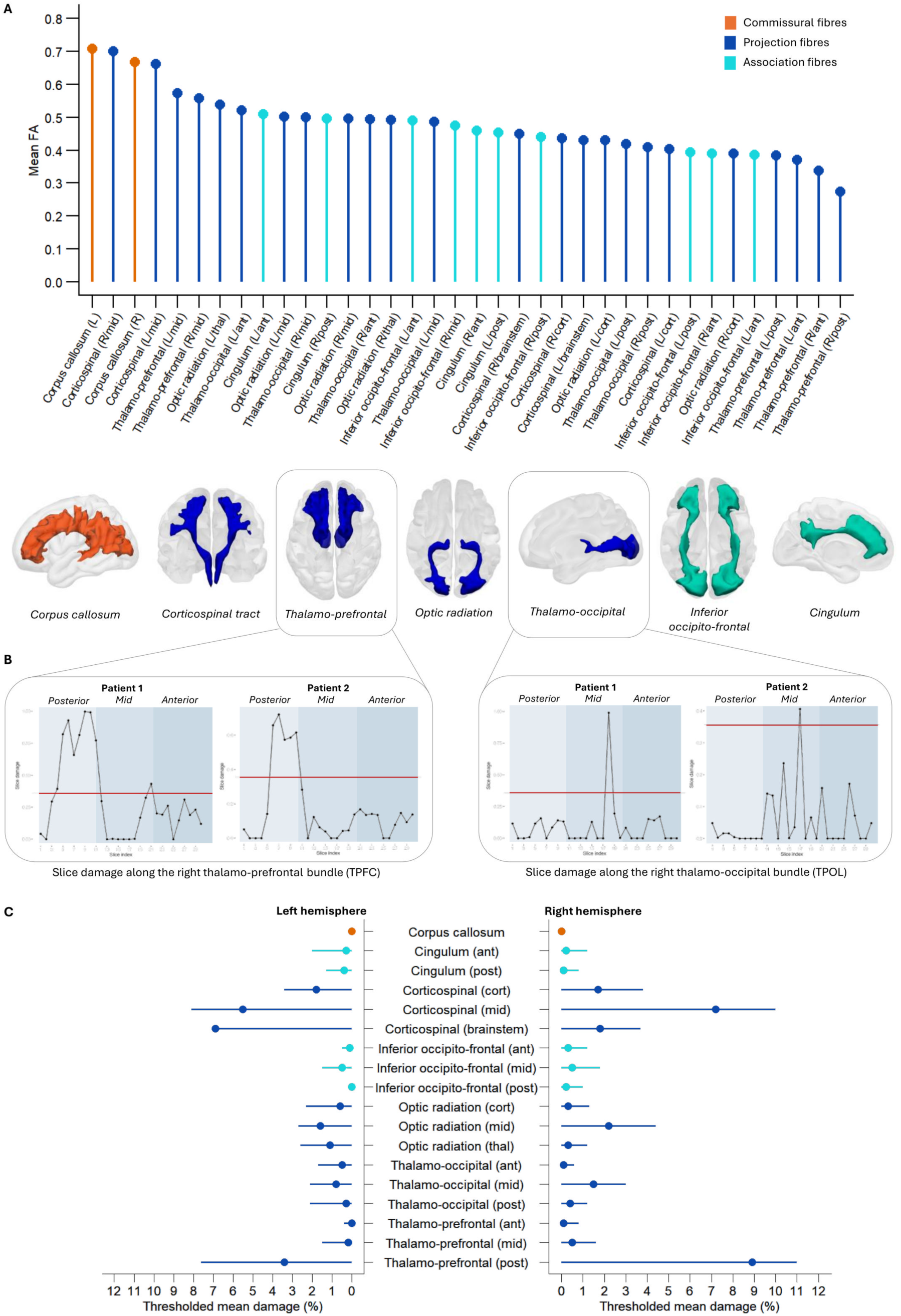
Diffusion-weighted imaging analysis in stable AQP4-IgG+NMOSD patients. **(A)** Mean fractional anisotropy (FA) of the white matter fibre tracts analysed in this study. **(B)** Patient-level examples of fibre damage along thalamus-adjacent white matter tracts. Patient 1 is a female in their 40’s with a history of two brainstem and myelitis attacks presenting with nausea, vertigo, double vision, paraesthesia, and leg weakness. Now in her “stable” phase, her acute symptoms have remitted completely (EDSS 1.5). Her MRI shows no remaining spinal lesions, but *n*=41 lesions in the brain parenchyma (volume: 1.6 mL). She continues to report severe fatigue (FSS 4.6). Patient 2 is also a female in their 40’s with two preceding complex attacks. She experienced bilateral optic neuritis, bladder dysfunction, hyperaesthesia, and pain. At the time of her first “stable” disease phase MRI, she presented with minimal disability (EDSS 2.0), no pain (PD-Q), and low brain lesion load (*n*=1, volume: 0.12 mL). **(C)** Thresholded mean damage of left and right white matter bundles regions (means and standard deviations). Abbreviations: ant – anterior region, cort – cortical region, EDSS – Expanded Disability Status Scale, FSS – Fatigue Severity Scale, mid – middle region, L – left, post – posterior region, PD-Q – PainDETECT questionnaire, R – right, thal – thalamic region.

In comparison to the left corpus callosum fibre tract, all analysed tracts and regions had lower mean FA values, although the right corpus callosum, and left and right mid-regions of the CST were very similar with mean differences of -0.04, -0.05, and -0.01, respectively. The mid-regions of the left and right TPFC did not differ in mean FA than the left corpus callosum with mean differences of –0.14 and –0.15 (Supplementary Fig. 3).

### Longitudinal white matter fibre-bundle changes

Group longitudinal changes in mean FAs from fibre-bundle regions were assessed from the first available DWI session and consecutively between each follow-up DWI and the first available scan. Since Cohen’s *d* was calculated for all mean differences between visits, effect sizes were considered “moderate to large” if the values were ≥ 0.5 and ≤ –0.5.^61^

#### Whole cohort white matter regions with decreasing FA

The following fibre-bundle regions showed decreasing mean FA over time with some fluctuations: the 1) left cortical region of the OR, 2) left posterior region of the TPOL, and the 3) left posterior region of the TPFC. Here, low FA values early in the “stable” phase significantly predicted both lower functional (MSFC, left posterior TPOL: *ρₛ*=0.67, *P_FDR_*=0.028) and cognitive outcomes at last follow-up (SDMT, left posterior TPOL: *ρₛ*=0.62, *P_FDR_*=0.012, left cortical OR: *ρₛ*=0.60, *P_FDR_*=0.018). All regions showed moderate effect sizes in mean FA differences over time, mostly between the last follow-up and the first available clinical session, although this could indicate a bias in patients with more MRI sessions (*n*=4). The left posterior TPFC showed a consistent decrease in mean FA between the third clinical visit and first clinical visit of the patient cohort, with a seeming recovery of FA (i.e. increase in mean FA) over further visits (Table 4, Supplementary Fig. 4A-C). Further investigation of increases in FA can be found in the Supplementary Results section.

**Table 4.** Cohen’s *d* values of fibre-bundle regional mean FA differences between visits.

| Bundle | Region | Visit comparisons (Cohen's *d*) |  |  |  |  |
| --- | --- | --- | --- | --- | --- | --- |
|  |  | Visit 2 –<br>Visit 1 | Visit 3 –<br>Visit 1 | Visit 4 –<br>Visit 1 | Visit 5 –<br>Visit 1 | Visit 6 –<br>Visit 1 |
| Corpus Callosum | Left | 0.03 | -0.13 | -0.12 | -0.16 | 0.21 |
|  | Right | 0.13 | -0.05 | -0.06 | -0.17 | 0.16 |
| Cingulum | Left Anterior | -0.03 | -0.05 | -0.11 | -0.13 | -0.01 |
|  | Left Posterior | 0.01 | -0.23 | -0.26 | -0.18 | 0.34 |
|  | Right Anterior | 0.06 | -0.08 | -0.06 | 0.00 | -0.28 |
|  | Right Posterior | 0.13 | -0.07 | 0.04 | 0.02 | 0.39 |
| Corticospinal Tract (CST) | Left Brainstem | 0.16 | 0.20 | 0.30 | 0.04 | **0.55** |
|  | Left Mid | -0.12 | 0.24 | 0.39 | 0.19 | **0.74** |
|  | Left Cortical | 0.02 | 0.05 | 0.03 | 0.09 | 0.31 |
|  | Right Brainstem | 0.09 | 0.16 | 0.13 | 0.16 | -0.23 |
|  | Right Mid | -0.23 | -0.22 | 0.11 | 0.03 | 0.45 |
|  | Right Cortical | -0.09 | -0.07 | 0.09 | 0.34 | 0.40 |
| Inferior Fronto-Occipital Fasciculus (IFOF) | Left Anterior | 0.27 | 0.16 | 0.19 | -0.27 | -0.12 |
|  | Left Mid | 0.28 | 0.16 | 0.05 | -0.06 | 0.32 |
|  | Left Posterior | 0.05 | -0.12 | -0.15 | -0.12 | 0.38 |
|  | Right Anterior | 0.35 | 0.09 | 0.29 | 0.09 | 0.32 |
|  | Right Mid | 0.20 | 0.10 | 0.22 | **0.50** | **1.07** |
|  | Right Posterior | 0.14 | -0.02 | 0.13 | 0.29 | **0.55** |
| Optic Radiation (OR) | Left Cortical | 0.07 | 0.00 | 0.19 | 0.14 | **-0.54** |
|  | Left Mid | -0.08 | -0.27 | -0.26 | -0.19 | 0.07 |
|  | Left Thalamic | 0.12 | -0.14 | -0.21 | -0.10 | -0.08 |
|  | Right Cortical | 0.10 | 0.03 | 0.20 | 0.14 | 0.23 |
|  | Right Mid | 0.15 | -0.05 | 0.10 | 0.18 | 0.28 |
|  | Right Thalamic | 0.15 | 0.04 | 0.10 | 0.44 | 0.21 |
| Thalamic Projection to Occipital Lobe (TPOL) | Left Anterior | 0.04 | -0.21 | -0.27 | -0.14 | 0.07 |
|  | Left Mid | -0.07 | -0.27 | -0.22 | -0.17 | 0.13 |
|  | Left Posterior | 0.07 | 0.01 | 0.18 | 0.15 | **-0.50** |
|  | Right Anterior | 0.16 | 0.04 | 0.12 | 0.48 | 0.12 |
|  | Right Mid | 0.14 | -0.05 | 0.09 | 0.22 | 0.24 |
|  | Right Posterior | 0.13 | 0.05 | 0.19 | 0.15 | 0.24 |
| Thalamic Projection to Prefrontal Cortex (TPFC) | Left Anterior | -0.02 | -0.08 | 0.00 | -0.09 | 0.02 |
|  | Left Mid | 0.00 | -0.10 | -0.01 | 0.02 | -0.21 |
|  | Left Posterior | -0.35 | **-0.65** | 0.04 | -0.19 | 0.31 |
|  | Right Anterior | -0.07 | -0.24 | -0.24 | -0.27 | -0.43 |
|  | Right Mid | 0.01 | -0.10 | -0.12 | 0.09 | 0.37 |
|  | Right Posterior | -0.31 | -0.39 | -0.17 | 0.20 | **0.55** |

#### Individual patient white matter regional fibre-bundle damage

Intra-individual longitudinal fibre-bundle regional damage was evaluated between each consecutive MRI session available per patient. Regions with enough damaged white matter streamlines to surpass the cohort SRD of 0.356 (i.e. at least 35.6% of streamlines in a slice) showing what is considered damaged slices of a tractogram were: the left and right CST at the mid (mean thresholded damage: 5.5% and 7.2%, respectively) and left brainstem (6.9%) regions, the right mid-region of the optic radiation (2.2%), and the left and right posterior region of the TPFC (3.4% and 8.9%, respectively; Fig. 3B-C). Other regions with damage had less than 2.2% of streamlines in the bundle regions affected (Supplementary Table 3). Due to the methodology for calculating damaged regions, increases in FA in each vertex of a streamline per bundle and region are not currently available as an output measure.

### Secondary analysis of the subgroup with higher-thalamic-volumes

Post-hoc analyses revealed that the volume increase was largely driven by the upper tertile of patients (mean volume ± *SD*: 9,322 ±383 mm³, 95% *CI*: 9,048 to 9,596 mm³, Fig. 4A). The thalamic volume estimates of the middle and lower tertiles (7,496 ±780 mm³, 95% *CI*: 7,159 to 7,833 mm³), in contrast, resembled those of healthy control participants (7,446 ±534 mm³, 95% *CI*: 7,235 to 7,657 mm³).

**Figure 4.**
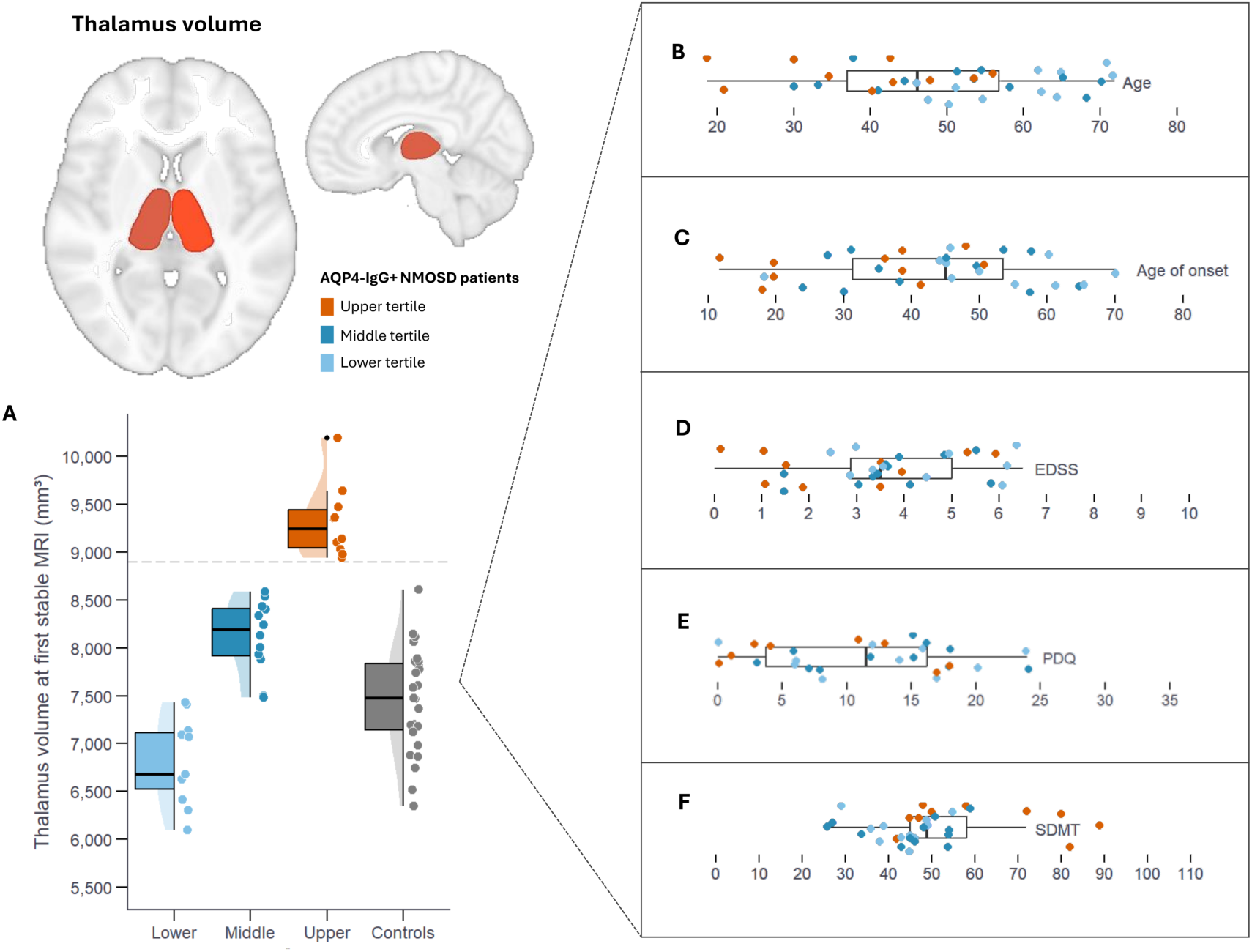
Thalamic volume subgroups in patients with “stable” AQP4-IgG+NMOSD. **(A)** Volumetry revealed that thalamic volumes are markedly increased in about 30% of the patients with relapse-free AQP4-IgG+NMOSD. Patients in the upper tertile of thalamic volumes were outside of the range of healthy participants. **(B-F)** A review of medical charts revealed that patients in the upper tertile of thalamic volume (orange) were typically younger, tended to have lower physical disability, often reported lower pain levels, and frequently had better cognitive performance. Abbreviations: EDSS – Expanded Disability Status Scale, PD-Q – PainDETECT Questionnaire, SDMT – Symbol Digit Modalities Test.

#### Clinical characteristics

Patients with high thalamic volumes were typically younger (upper thalamic tertile: median 41.4 [*IQR*: 31.3-46.8] years, middle and lower tertiles: 54.5 [46.9-64.6] years), had a younger age of onset (37.6 [19.8-40.6] vs. 45.8 [36.7-57.6] years), tended to have lower physical disability (EDSS, 2.8 [1.1-3.9] vs. 3.5 [3.1-5.0]), reported less pain (PD-Q sum score: 3.5 [0.3-12.5] vs. 13.0 [6.3-16.8]), and performed better on cognitive tests (e.g., SDMT: 61.3 ±17.7 vs. 44.1 ±9.1). Moreover, they were generally less likely to have comorbidities (10% vs. 74%), and 60% of them were on attack-preventing treatment (rituximab). In contrast, disease courses were largely comparable across subgroups, with similar disease duration (5.7 [2.9-9.9] vs. 5.3 [3.3-8.1] years), time since last attack (1.8 [1.7-2.5] vs. 1.8 [1.6-3.1] years), and number of attacks (3.0 [2.0-4.0] vs. 3.0 [2.0-4.5]). Rates of myelitis (historically: 100% vs. 91%, as last attack: 60% vs. 57%) and optic neuritis (historically: 70% vs. 57%, as last attack: 30% vs. 35 %) were also comparable.

#### White matter profiles of fibre-bundle regions

Compared to the rest of the cohort, these ten patients showed large effect sizes for the mean difference in FA values of several thalamus-adjacent white matter regions in the first DWI (Supplementary Table 4).These regions were the cingulum (left anterior and left and right posterior regions), IFOF (right anterior region), TPOL (left mid region), IFOF (right anterior region), and the TPFC (right anterior and posterior regions). In all these regions, the patients with upper tertile thalamic volumes showed higher mean FA values (mean differences between –0.02 to –0.05; Cohen’s *d* between 0.82 to 1.33) than the rest of the cohort (Supplementary Fig. 5).

We further evaluated two patients with the largest thalamic volumes for their individual regional fibre-bundle damage profiles over time. These patients showed microstructural damage over each consecutive DWI scan in every region of the CST, with the highest amount in the mid-regions. Both patients also showed a high amount of damage in the right posterior region of the TPFC and a moderate amount of damage in the right mid-region of the TPOL (Fig. 3B). In the patient with the highest thalamic volume, there was additional damage in the left mid-region of the OR. Meanwhile, the other patient had more widespread and higher damage values, including in the left posterior and right anterior cingulum, right posterior IFOF, right mid-region of the OR, and the left posterior region of the TPFC. Although we could not evaluate increases in FA in fine detail using Gaussian mixture models to identify specific vertices where this may occur, we were able to analyse the mean FA in each tract region between the first and second MRI sessions. These two representative patients showed 16 regions with a mean increase in FA of 2.11% (± 0.64%) over time (Supplementary Table 5), with three of them overlapping (right mid-region of the IFOF, right thalamic region of the OR, and right anterior region of the TPOL).

## Discussion

This study addressed the hypothesis that subtle volumetric and white matter microstructural changes occur in “stable” AQP4-IgG+NMOSD. We investigated this by measuring brain volumes and white matter integrity with MRI and then related these metrics longitudinally to clinical symptoms and cognitive performance. Several novel MRI-related changes were observed over time: 1) Cortical grey matter atrophy occurred at an increased rate compared to normal healthy ageing. 2) Thalamic volumes were increased compared to healthy participants, especially in a subgroup with a mild disease course prior to this “stable” phase. 3) Several white matter fibre-bundles showed group-level decreases in structural integrity, including the optic radiation and thalamic projections to the occipital lobe. Longitudinally, the most common regional white matter damage tracked over time was found in the mid-region of the CST, the posterior region of the thalamic projection to the prefrontal cortex, and the mid-region of the optic radiation.

Our volumetric approach revealed that global brain atrophy was mainly driven by grey matter – specifically cortical – volume loss over time. This atrophy exceeded normal ageing rates found in large-scale reference curves (*Braincharts*) and was related to worse motor outcomes and higher self-reported current pain levels at last follow-up. These volumetric findings are in line with a recent meta-analysis reporting moderate to large overarching effects for whole brain and grey matter volume in patients with AQP4-IgG+NMOSD.^24^ Here, smaller brain volumes were associated with lower cognitive scores on the PASAT and more previous attacks. Interestingly, longitudinal brain volume loss in the relapse-free phase has also been found to associate with increased serum glial fibrillary acidic protein, a marker for astrocytic damage.^4^ Cortical grey matter atrophy is less pronounced in patients with AQP4-IgG+NMOSD than in multiple sclerosis,^28^ with regional atrophy most likely occurring in the occipital cortex, frontal areas, and the cerebellum.^24,28^ Recent studies have found that grey matter reductions in NMOSD related to worse performance on tests of executive function^30^ and memory,^62^ while subcortical grey matter volumes appear to be frequently spared.^25,28,63,64^ However, decreased deep grey matter volumes can be observed for a subgroup of patients with cognitive impairment, including the thalamus, caudate, and hippocampus.^20,65^

About a third of the patients experience cognitive impairment, making it a notable and potentially overlooked symptom.^20,30^ Attention, processing speed, and memory are frequently affected.^20,66^ Here, we found that thalamic volumes were, on the group level, *higher* compared to healthy participants. Similarly, pallidum volumes increased significantly over time. Higher thalamic volumes were mainly driven by patients who were younger, had fewer historic attacks, and few comorbidities. Patients with higher initial thalamic volumes also performed better on tests of attention (SDMT) and spatial memory (SPART) later in this study. This finding challenges the current understanding of the role of the thalamus in NMOSD. How thalamic volumes are affected likely depends on the disease phase (i.e., relapse vs. relapse-free), whether specific subgroups are analysed (e.g., with and without cognitive impairment), and the observation period (i.e., cross-sectional or longitudinal designs). Aligned with this proposition, there were no overarching significant effects in either direction in a volumetric meta-analysis.^24^ Additional studies are required to determine the role of ageing,^67,68^ sex-specific influences pertaining to inflammation and neurodegeneration,^69^ as well as potential beneficial roles of brain and cognitive reserve^70^ in thalamic and basal ganglia neuroplasticity. As the choroid plexus has been shown to release inflammatory mediators into the brain, it is interesting to study in a relapse-free phase of the NMOSD disease.^71^ We did not observe any change in the choroid plexus volumes over time. Previously, a large study showed no difference in choroid plexus volumes in patients with NMOSD versus healthy participants.^72^ Thus, it would not be expected to differ, especially in “stable” patients. Although one recent study found MRI intensity and texture differences of the NMOSD choroid plexus, there were no shape differences compared to healthy participants. While this points to a possible involvement in the disease,^73^ our volumetric study method may not be sensitive enough to detect any changes in the relapse-free disease phase.

White matter affection in NMOSD is a secondary effect of initial astrocytopathy by acute attacks.^15^ ^74–76^ From our analyses, we could see that several regions were highly impacted in AQP4-IgG+NMOSD, despite a relatively “stable” disease course. At a cross-sectional level, the first available DWI scans showed widespread low mean FA in most of the white matter fibre-bundles we investigated, although not out of the normal range in the cohort age-group. However, when evaluating the longitudinal changes in mean FA, we could observe that three main regions showed overall decreases in white matter integrity: the right posterior region of the TPFC, the mid- and brainstem regions of the CST, and the right mid-region of the optic radiation. Two of these regions are related to the visual pathway in the brain.^74,75^ Here, white matter integrity loss in the posterior TPFC was associated with functional and cognitive outcomes, including higher general disability, worse motor function, and worse performance on tests of higher attention, visuospatial, and verbal memory. Additionally, decreased white matter integrity in the left posterior TPOL and left cortical region of the OR early in the “stable” phase predicted both lower functional and cognitive outcomes at last follow-up.

Interestingly, all regions are relatively close to the thalamus. The mid-region of the CST has fibres that run through the internal capsule, which lies between the thalamus and the lentiform nucleus. Previous studies in stroke and spinal cord injury have shown that damage to the white matter in the internal capsule region leads to worse motor outcomes and upstream damage occurs within the first few months of cord injury with relatively fast degeneration.^76,77^ In the region of the brainstem, CST white matter damage is associated with weakness and paralysis.^78,79^ These symptoms and regions are undoubtedly important to assess in NMOSD, as evidenced in our relapse-free cohort, where a high percentage of streamlines in the white matter fibre bundle regions were damaged even in the patients with the highest thalamic volumes. The other white matter fibre-bundle regions showing longitudinal microstructural “damage” in the patients with high thalamic volumes are both directly connected to the thalamus - the optic radiation to the lateral geniculate nucleus and the TPFC to the medial dorsal nucleus - with one directly involved in visual sensory processing and the other in higher-order cognition.^80,81^

Our longitudinal analysis of white matter fibre-bundle regions gave interesting results, both on the group level and in the representative patients with higher thalamic volumes. While many white matter tracts showed microstructural damage, the regions closest to the thalamus in the IFOF, OR, and TPOL showed slight increases in mean FA over time in the entire cohort without thresholding. FA increases could be thought of as white matter regeneration, however, these minute increases (<5%) could be within normal fluctuation of the measurement between patient timepoints.^82^ All tracts and regions would likely show “damage” over time, where normal ageing contributes to integrity decline based on myelin deterioration.^83^ Whether there is increased myelin deterioration during the “stable” disease phase is not clear. However, it has been found that association fibres, such as the IFOF and OR, are less likely to show a decrease in FA due to ageing only. Even in adults, there is heterogeneity in white matter integrity loss in ageing in the projection fibres, such as the CST.^84^ The representative patients we evaluated only had two timepoints available for analyses. Again, in the two representative patients, the overlapping regions showing increased FA were directly beside or connected to the thalamus.

There are several limitations that need to be considered. Even though the intervals between the yearly follow-up visits were mostly evenly distributed, there is some variation. At the same time, some patients had more visits than others. To account for this, we used longitudinal processing approaches^43^ and applied linear mixed effects models that are deemed most adequate to handle variability and potential missing data points in longitudinal designs. While most patients are diagnosed in midlife, the age of onset can range substantially (11.7-70.1 years in our sample). These potential developmental and ageing effects need to be taken into account. Our approach to model participants as random intercept is a strength of our study, as it can help address the fact that individual patients may have started out at different levels and, consequently, their potential for change may vary.^90^ However, we cannot fully exclude that atrophy rates differ based on when patients enter the “stable” phase. Second, a common potential confounder in longitudinal cognitive testing are practice effects, particularly for shorter retest intervals.^91^ Here, better scores may reflect repeated exposure to the testing material and more familiarity with the task rather than a true, underlying improvement. While this study applied common measures to limit practice effects, such as longer retest intervals and using parallel versions where applicable, we cannot fully exclude them.

It should be noted that since we could not calculate the number of streamlines with a true “increase” in FA (greater than the SRD), findings related to increased FA likely would not surpass a threshold for true difference between DWI imaging timepoints. Nonetheless, longer follow-ups may reveal myelin repair in some regions, however rare. When evaluating the regional damage over time using a more sensitive, intra-individual fibre-bundle region longitudinal tracking method (Mewes et al., in preparation), we found that the CST and TPFC showed a steady decrease. As higher false-positives may be found in evaluating hundreds of FA streamlines and averaging them across individuals and timepoints, few conclusions can be made for such a rare heterogeneous disease. Our current intra-individual fibre-bundle regional analysis method, including the calculation and thresholding of true “damage” based on the smallest real difference is a strength of our study, allowing for disentangling the age- versus disease-related longitudinal decreases in white matter integrity. Thus, in future studies, we would propose using a method for individual evaluation of subtle white matter integrity changes in AQP4-IgG+NMOSD.

Lastly, even though we harnessed the potential of large-normative volumetric reference scores, there was no longitudinal, age-matched MRI data from healthy participants available at our centre. Braincharts models sex- and age-stratified non-linear growth trajectories over the entire lifespan.^45^ In Braincharts’ longitudinal subset, a proof-of-concept analysis revealed that volumetric centile scores had low within-subject variability across repeated assessments. This demonstrates that centile scores in individuals are generally stable over time, providing a basis for their use in longitudinal designs. However, not all regional volumes of interest in AQP4-IgG+NMOSD are represented in Braincharts. This highlights the need for future research on regional brain health in AQP4-IgG+NMOSD, ideally down to the subfield level.

## Conclusion

This in-depth regional brain volumetric and microstructural integrity analysis in patients with a longitudinal “stable” disease course of AQP4-IgG+NMOSD resulted in intriguing new insights into possible thalamic plasticity with white matter damage and repair profiles. These imaging findings point to a better clinical and cognitive prognosis for younger patients with few or no comorbidities.

## Data availability

Due to ethical and privacy restrictions protecting participant confidentiality, the data are not publicly available but can be obtained from the corresponding author upon reasonable request and subject to institutional approval.

## Acknowledgements

We would like to thank the Berlin Center of Advanced Imaging and Neuroscience Clinical Research Center for their support in MRI and clinical data collection and administration, as well as Susan Pikol and Cynthia Kraut for their expert brain lesion segmentation and MRI scanning of patients and healthy participants. Finally, we would like to thank all the patients and healthy participants for their time and participation in support of this study.

