## Supplementary Methods for "Longitudinal advanced MRI changes in relapse-free patients with AQP4-IgG+NMOSD"

#### MRI data acquisition

MRI images were acquired on a 3T Siemens MAGNETOM Tim Trio MRI machine (Siemens, Erlangen) equipped with a 32-channel head coil at the Berlin Center of Advanced Neuroimaging (BCAN). The imaging protocol included a 3D 1mm-isotropic T1 weighted magnetization prepared rapid acquisition gradient echo (MPRAGE) sequence (TR/TE/TI=1900/2.55/900 ms, matrix 240×240[PCC51.1]), a 3D 1mm-isotropic fluid-attenuated inversion recovery sequence (3D FLAIR) (TR/TE/TI=6000/388/2100 ms, matrix=256×256), a single-shot echo planar imaging diffusion-weighted imaging (DWI) sequence (TR/TE=7500/86 ms, matrix 96×96, 61 slices no gap, slice thickness 2.3 mm, 64 non-colinear directions, b-value=1000 s/mm$^2$), as well as a 2D-sagittal T2-weighted SC sequence (slice thickness=2 mm, TR=3500 ms, TE=101 ms, in-plane resolution=0.91 mm×0.91 mm) at the cervical, thoracic, and lumbar level of the spinal cord. Follow-up assessments were conducted using an identical imaging protocol and test battery across all time points. Comorbidities were recorded based on Samadzadeh et al..[1]

#### Brain T2-weighted lesion segmentation

The eight categories were: 1) corpus callosum (>50% coverage in length), 2) lateral ventricle, 3) brainstem including fourth ventricle, 4) cerebellum, 5) juxtacortical, 6) diencephalic including third ventricle, 7) corticospinal tract extending into the pons, and 8) non-specific white matter lesions.[2] Brain total T2-hyperintense lesions were counted and volumes extracted in mL using FSL cluster (https://fsl.fmrib.ox.ac.uk/fsl/docs/statistics/cluster.html?h=cluster).

#### Longitudinal fibre-bundle regional damage calculation

For calculating the number of slices per bundle region, the median coordinates of streamline endpoints in start and end regions defined in the tractogram *TractSeg* output were calculated for each bundle in R. Tractograms for the bundles of interest were then resampled to equidistant slices in a straight line between the median bundle starting and endpoints using MRtrix3 *tckresample*. The number of slices per bundle was selected by dividing the distances between starting and endpoints per tractogram by the largest dimension of the DWI voxel-size (2.5cm) and rounding to the closest multiple of 10. This was chosen, as the constraints of MRI resolution should be taken into consideration when creating slices to measure from in a tractogram. This total number of slices sampled were further subdivided into two to three regions depending on the tract of interest. For example, the CST (left and right) had 50 sampled slices, which is not a multiple of three. Thus, to achieve three regions, it was decided to select the first 15 slices as the cortical projecting region, the next 20 slices as the mid-region, and the last 15 slices as the brainstem projecting region. When the total number of slices were a multiple of two or three, all regions contained equal number of slices. In the case of the corpus callosum, the total number of slices were only subdivided into left and right hemispheres (five slices per hemisphere). The other WM tracts of interest analysed were: IFOF with three regions (anterior = 15 slices, mid = 10 slices, posterior = 15 slices); cingulum with three regions (anterior = 15 slices, posterior = 15 slices); optic radiation with three regions (cortical = 10 slices, mid = 10 slices, thalamic = 10 slices); TPOL with three regions (anterior = 10 slices, mid = 10 slices, posterior = 10 slices); and TPFC with three regions (anterior = 10 slices, mid = 10 slices, posterior = 10 slices).

### Supplementary Results

#### Whole cohort white matter regions with increasing FA

Meanwhile, moderate to large effect sizes were found for increasing mean FA longitudinally in the following regions with some fluctuations: the 1) left brainstem region of the CST, 2) left mid region of the CST, 3) right mid region of the IFOF, 4) right posterior region of the IFOF, and the 5) right posterior region of the TPFC. Over time, greater FA increases in the CST were significantly associated with better attention (PASAT 3s: $\beta$=0.002, $SE$=0.0002, $P_{FDR}$=0.017) and spatial memory outcomes at last visit (SPART delayed recall: $\beta$=0.011, $SE$=0.001, $P_{FDR}$=0.017, Supplementary Fig. 2). Similarly, higher FA gains in the right mid IFOF predicted better functional outcomes (MSFC: $\rho_s$=0.68, $P_{FDR}$=0.026) and better self-rated visual function at last follow-up (VFQ25: $\rho_s$=0.64, $P_{FDR}$=0.011). All regions showed the largest increase in FA in the comparison between the last follow-up and the first clinical visit MRI, again with the possibility that the data is skewed towards patients with more visits (*n*=4). However, the left brainstem and left mid-regions of the CST and the right mid and right posterior regions of the IFOF showed consistent increasing mean FA over all sessions. Meanwhile, the right posterior TPFC showed an interesting decrease in mean FA between the third MRI visit and first available clinical visit (*n*=27), then showed increased mean FAs compared to the first DWI scans (Table 4, Supplementary Fig. 4D-H).

---

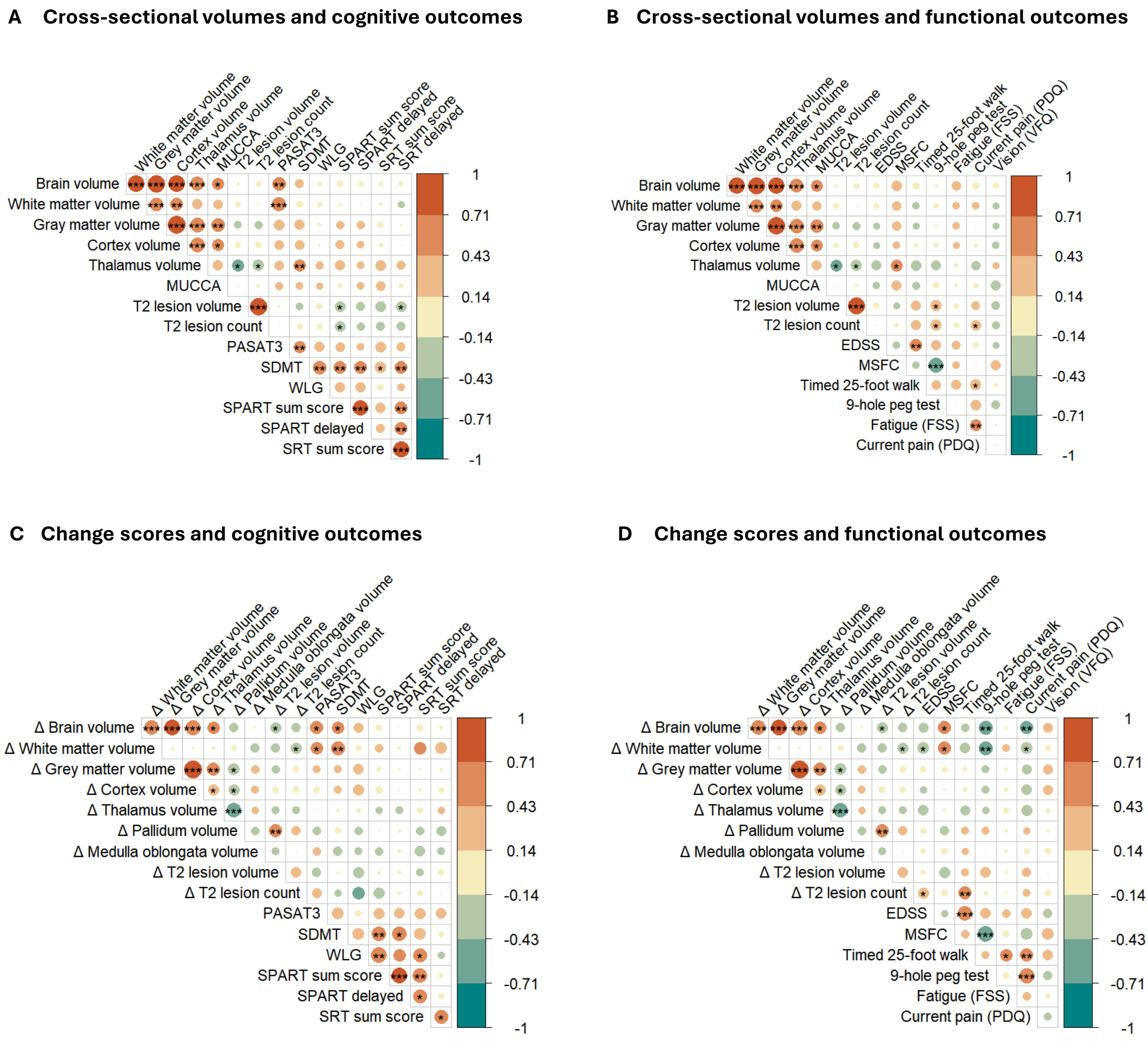


**Fig. S1. Visualization of Spearman correlation plots showing initial associations at first stable visit (A-B), and between longitudinal change and outcomes at last visit (C-D).** Abbreviations: EDSS – Expanded Disability Status Scale, FSS – Fatigue Severity Scale, MSFC – Multiple Sclerosis Functional Composite, MUCCA – Mean Upper Cervical Cord Area, PASAT – Paced Auditory Serial Addition Test, PDQ – PainDETECT Questionnaire, SDMT – Symbol Digit Modalities Test, SPART – Spatial Recall Test, SRT – Selective Reminding Test, VFQ – Visual Function Questionnaire 25, WLG – Word List Generation. (* $P_{unadj} < 0.05$, ** $P_{unadj} < 0.01$, *** $P_{unadj} < 0.001$)

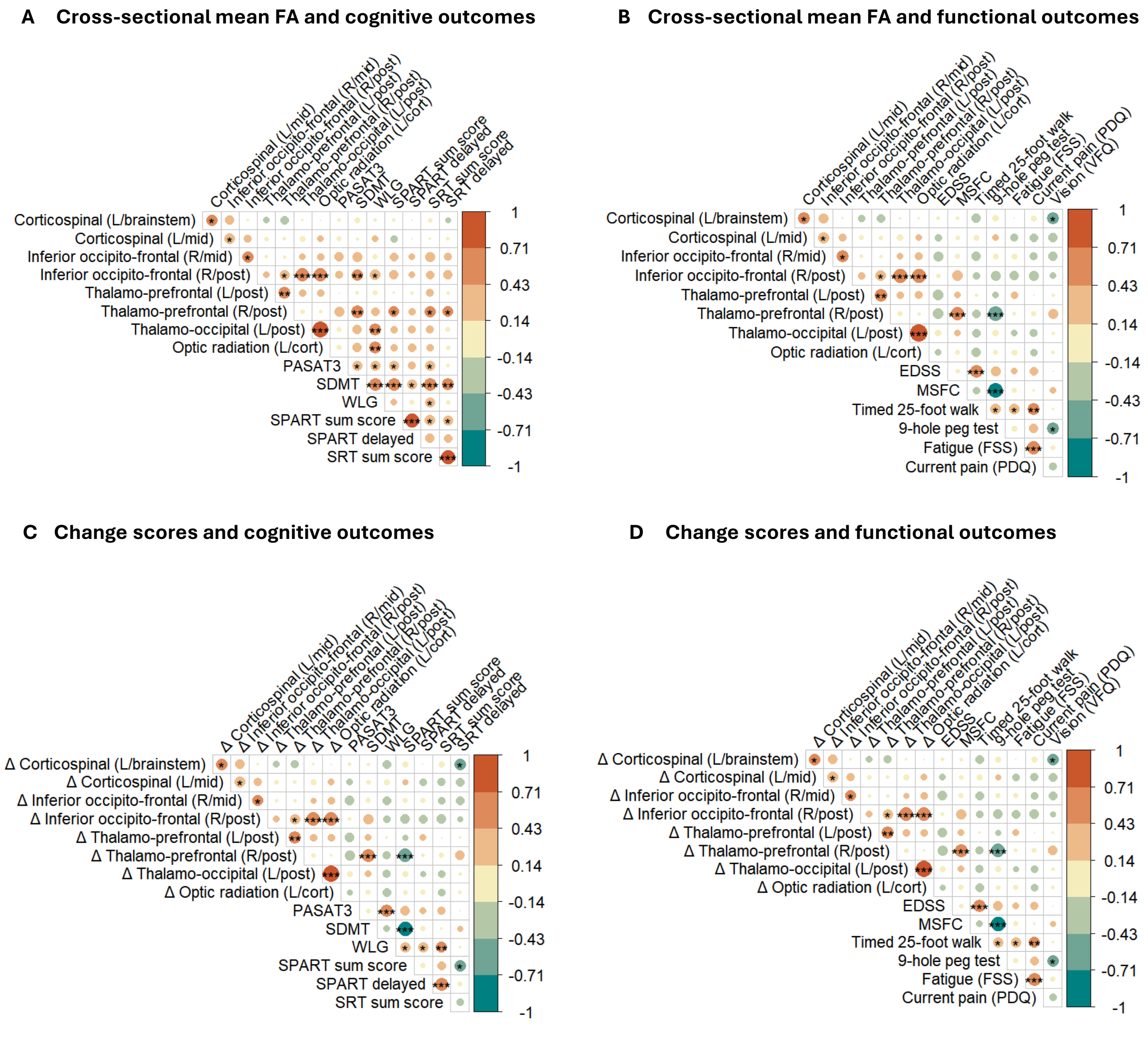


**Fig. S2. Visualization of Spearman correlation plots showing initial associations at first stable visit (A-B), and between longitudinal change and outcomes at last visit (C-D).** Abbreviations: cort – cortical, EDSS – Expanded Disability Status Scale, FA – Fractional Anisotropy, FSS – Fatigue Severity Scale, L – left, MSFC – Multiple Sclerosis Functional Composite, MUCCA – Mean Upper Cervical Cord Area, PASAT – Paced Auditory Serial Addition Test, PDQ – PainDETECT Questionnaire, post – posterior, R – right, SDMT – Symbol Digit Modalities Test, SPART – Spatial Recall Test, SRT – Selective Reminding Test, VFQ – Visual Function Questionnaire 25, WLG – Word List Generation. (* $P_{unadj} < 0.05$, ** $P_{unadj} < 0.01$, *** $P_{unadj} < 0.001$)

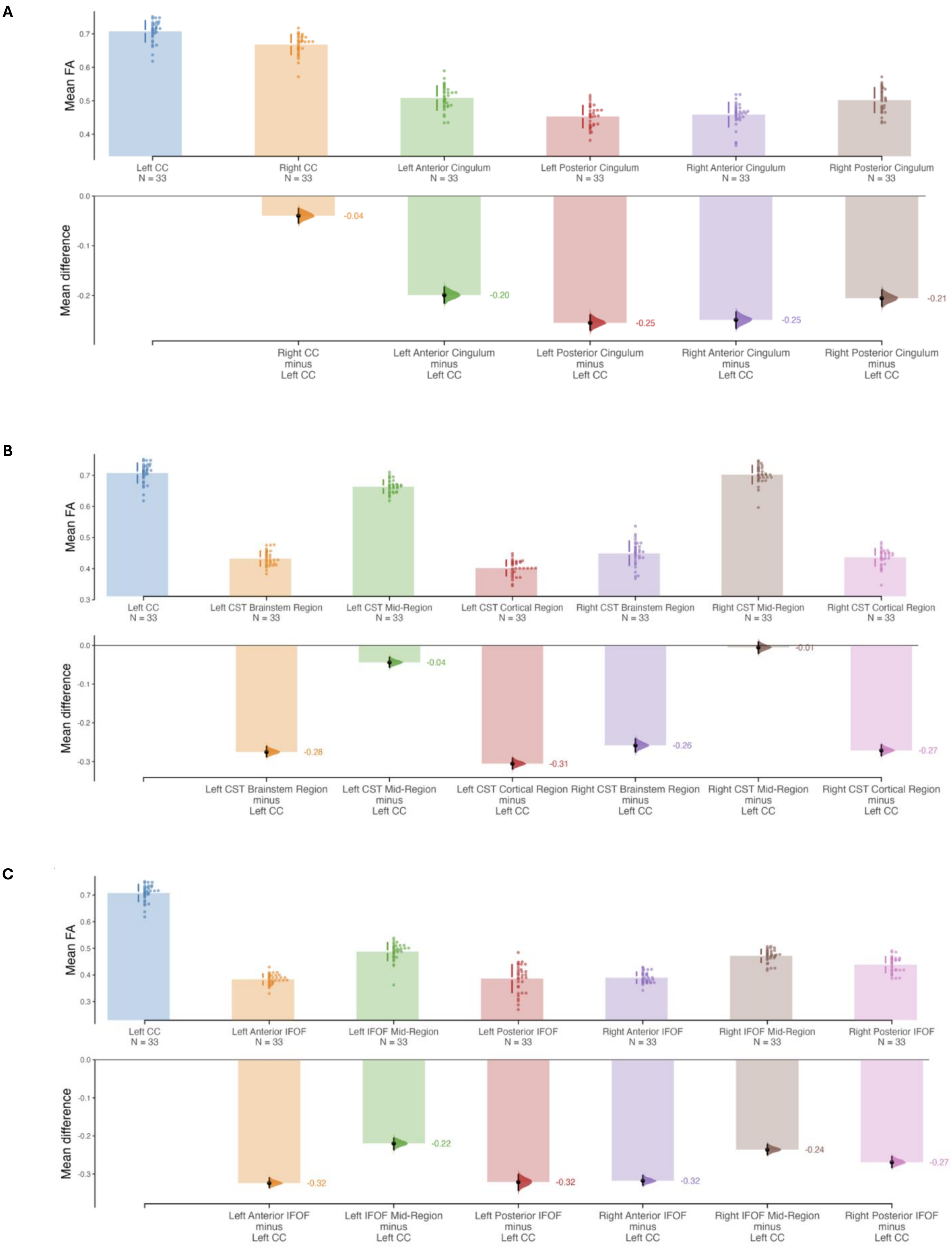


**Fig. S3. Mean differences in regional mean FA measures between the left CC and the rest of the WM fibre bundle regions.** Between the **(A)** left CC, right CC, and cingulum regions; **(B)** left CC and CST regions; **(C)** left CC and IFOF regions; **(D)** left CC and OR regions; **(E)** left CC and TPFC regions; and **(F)** left CC and TPOL regions. Abbreviations: CC – corpus callosum, CST – corticospinal tract, FA – fractional anisotropy, IFOF – inferior fronto-occipital fasciculus, OR – optic radiation, TPFC – thalamic projection to the prefrontal cortex, TPOL – thalamic projection to the occipital lobe, WM – white matter.

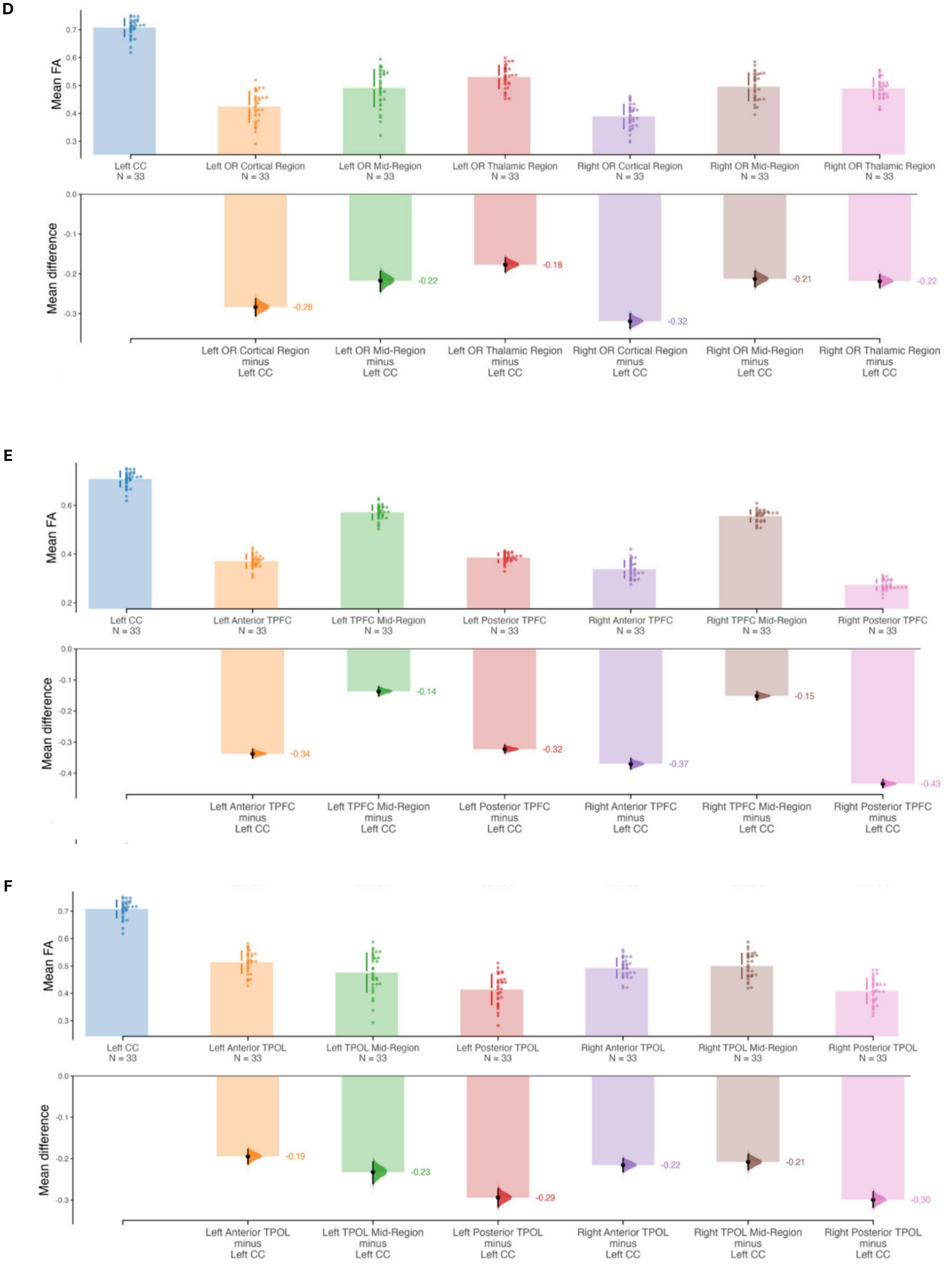


**Fig. S3. Mean differences in regional mean FA measures between the left CC and the rest of the WM fibre bundle regions.** Between the **(A)** left CC, right CC, and cingulum regions; **(B)** left CC and CST regions; **(C)** left CC and IFOF regions; **(D)** left CC and OR regions; **(E)** left CC and TPFC regions; and **(F)** left CC and TPOL regions. Abbreviations: CC – corpus callosum, CST – corticospinal tract, FA – fractional anisotropy, IFOF – inferior fronto-occipital fasciculus, OR – optic radiation, TPFC – thalamic projection to the prefrontal cortex, TPOL – thalamic projection to the occipital lobe, WM – white matter.

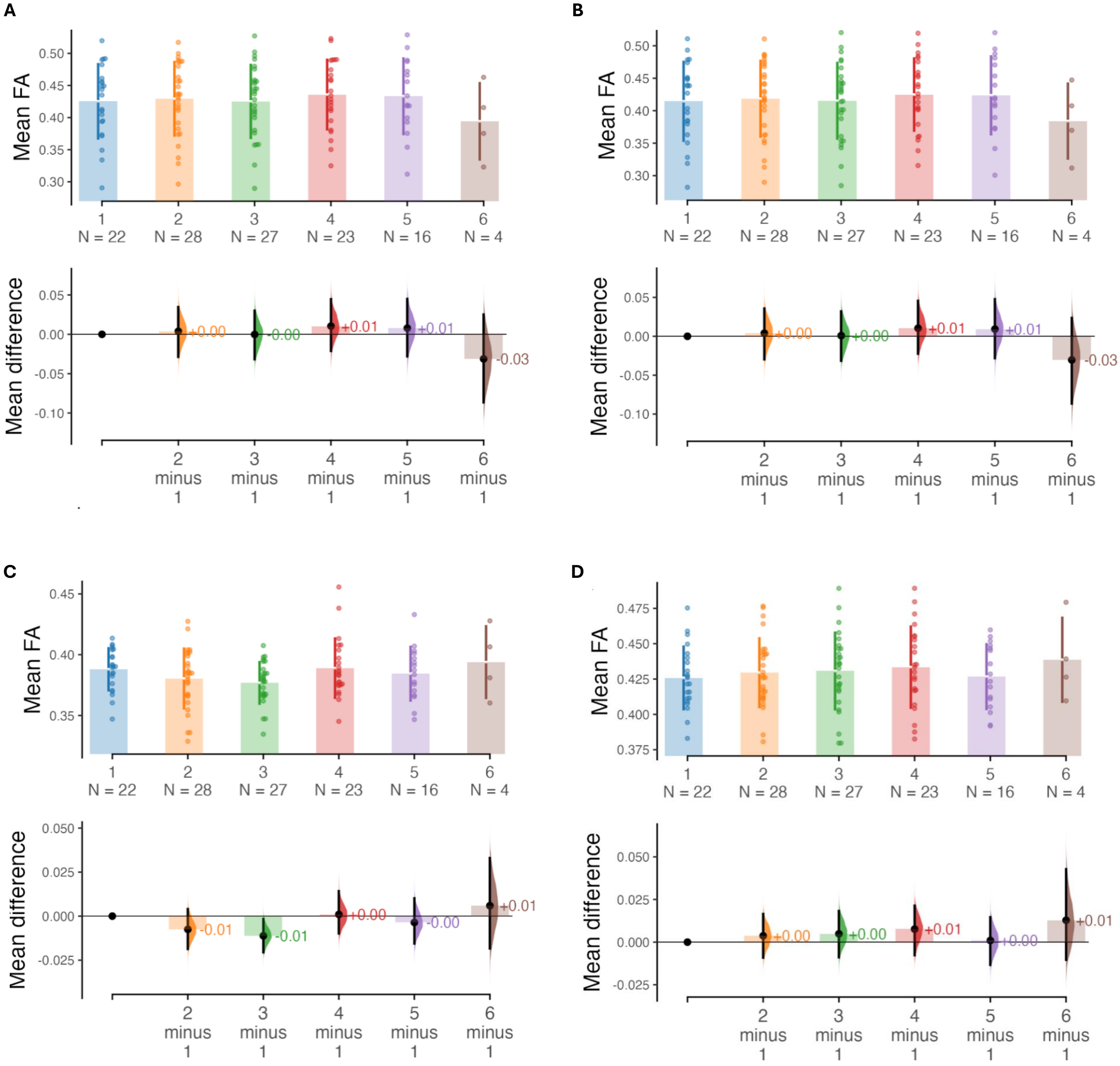


**Fig. S4. Whole cohort mean differences between mean FA values calculated at each available DWI timepoint in regions showing decreased and increased FA.** Mean differences were calculated between each follow-up timepoint and the first available scan showing decreasing FAs in the: **(A)** left cortical region of the OR; **(B)** left posterior region of the TPOL; and **(C)** left posterior region of the TPFC. Meanwhile, mean differences calculated between each follow-up timepoint and the first available scan showed increasing FAs in the: **(D)** left brainstem region of the CST; **(E)** left mid-region of the CST; **(F)** right mid-region of the IFOF; **(G)** right posterior region of the IFOF; and **(H)** right posterior region of the TPFC. Abbreviations: CST – corticospinal tract, DWI – diffusion weighted imaging, FA – fractional anisotropy, IFOF – inferior fronto-occipital fasciculus, OR – optic radiation, , TPFC – thalamic projection to the prefrontal cortex, TPOL – thalamic projection to the occipital lobe.

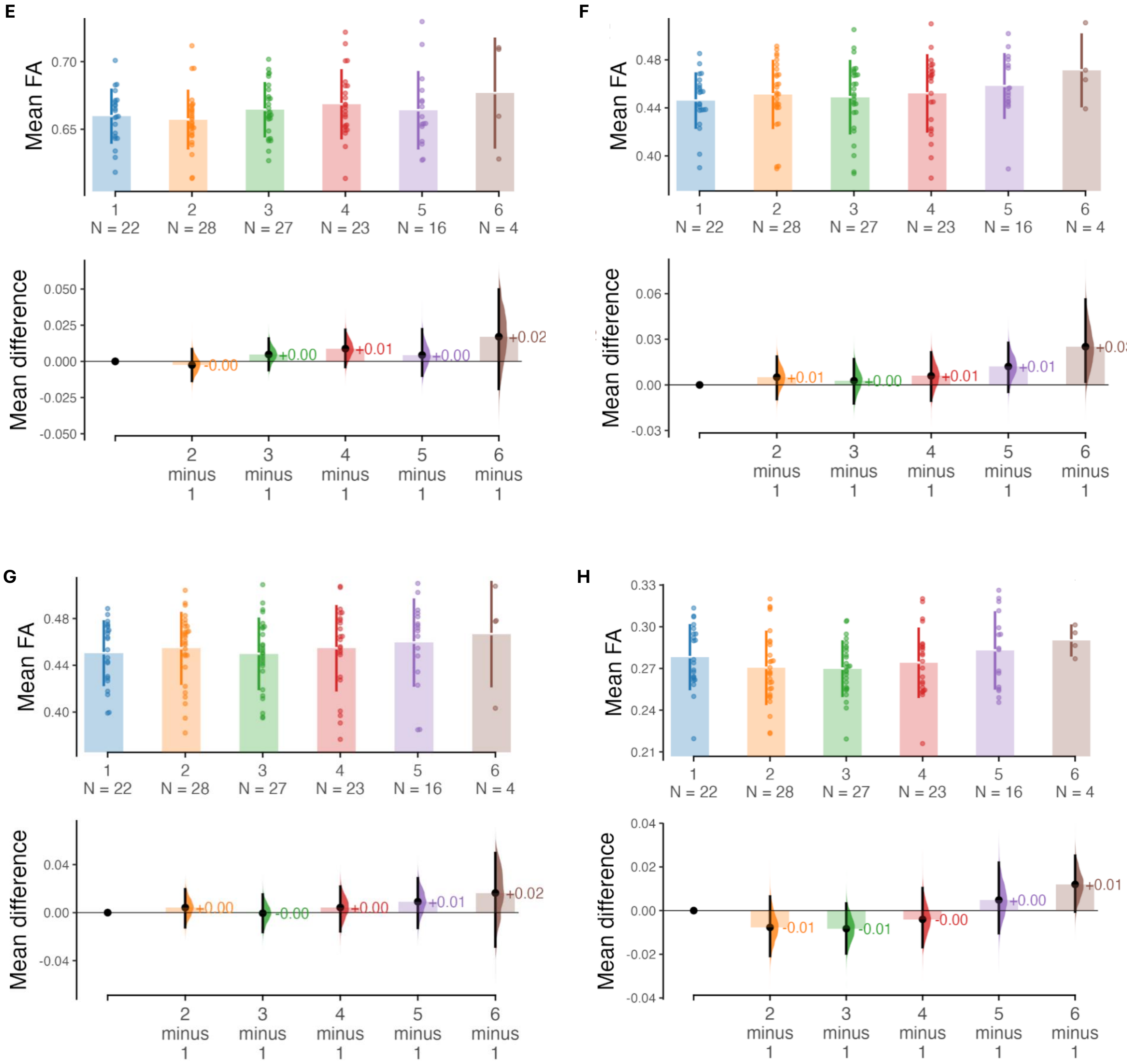


**Fig. S4. Whole cohort mean differences between mean FA values calculated at each available DWI timepoint in regions showing decreased and increased FA.** Mean differences were calculated between each follow-up timepoint and the first available scan showing decreasing FAs in the: **(A)** left cortical region of the OR; **(B)** left posterior region of the TPOL; and **(C)** left posterior region of the TPFC. Meanwhile, mean differences calculated between each follow-up timepoint and the first available scan showed increasing FAs in the: **(D)** left brainstem region of the CST; **(E)** left mid-region of the CST; **(F)** right mid-region of the IFOF; **(G)** right posterior region of the IFOF; and **(H)** right posterior region of the TPFC. Abbreviations: CST – corticospinal tract, DWI – diffusion weighted imaging, FA – fractional anisotropy, IFOF – inferior fronto-occipital fasciculus, OR – optic radiation, , TPFC – thalamic projection to the prefrontal cortex, TPOL – thalamic projection to the occipital lobe.

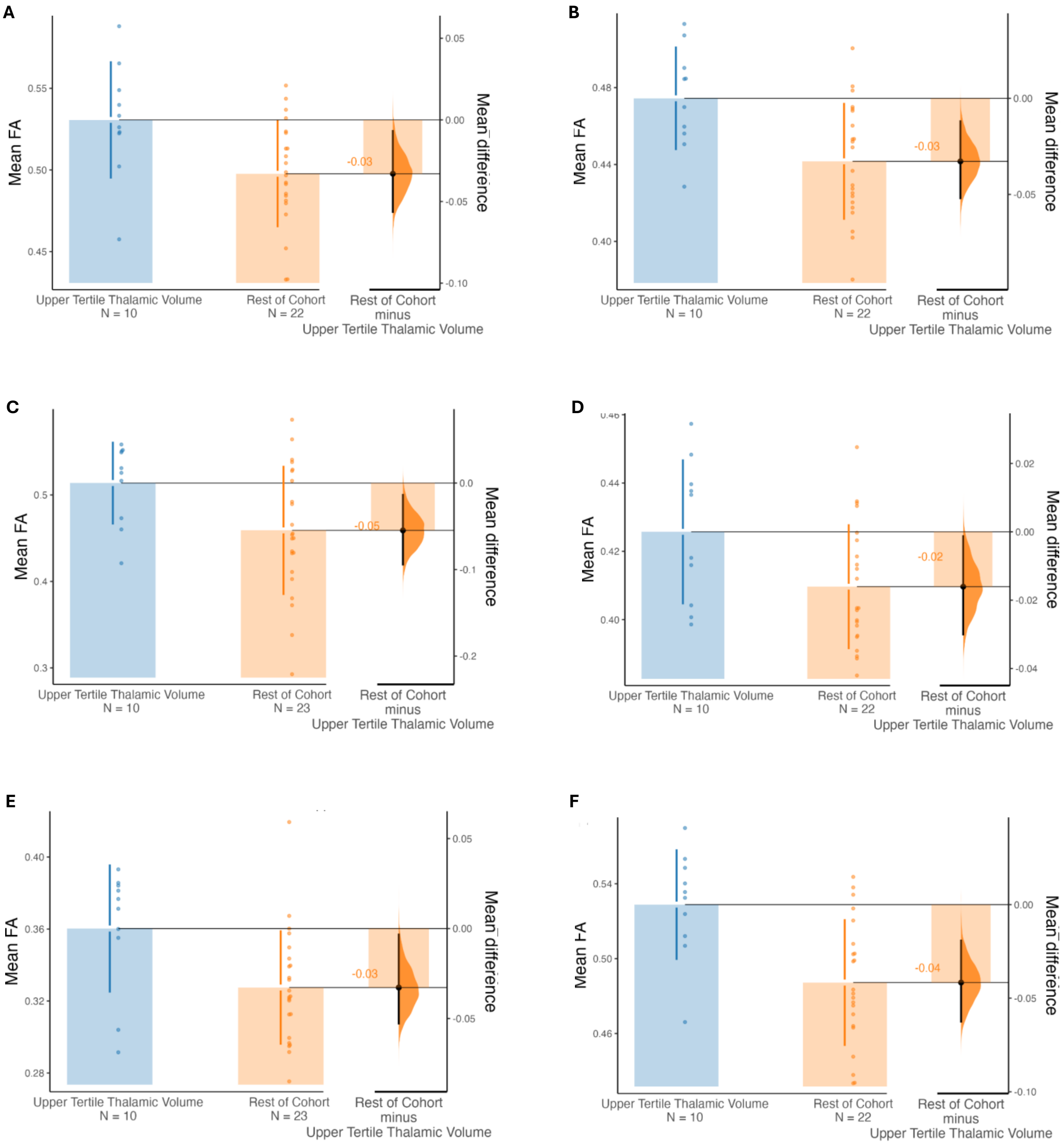


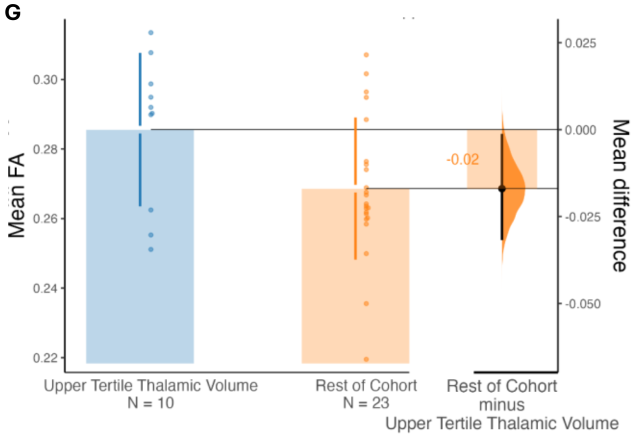


**Fig. S5. Mean differences between mean FA values calculated in the first available DWI scan in patients within the upper tertile of thalamic volumes and the rest of the cohort.** Regions shown: **(A)** Left Anterior Cingulum; **(B)** Left Posterior Cingulum; **(C)** Left TPOL Mid-Region; **(D)** Right Anterior IFOF; **(E)** Right Anterior TPFC; **(F)** Right Posterior Cingulum; **(G)** Right Posterior TPFC. Abbreviations: DWI – diffusion weighted imaging, FA – fractional anisotropy, IFOF – inferior fronto-occipital fasciculus, TPFC – thalamic projection to the prefrontal cortex, TPOL – thalamic projection to the occipital lobe.

**Supplementary Table 1 | Treatments, comorbidities, and study exclusion**

| | N (%) | Comment |
|---|---|---|
| **Comorbidities** | | |
| Patients with any comorbidity | 18/33 (55%) | |
| Autoimmune (any) | 17/33 (52%) | |
| Endocrine (any) | 8/33 (24%) | |
| Rheumatologic (any) | 7/33 (21%) | |
| Thyroid disorder | 6/33 (18%) | |
| Systemic lupus erythematosus (SLE) | 3/33 (9%) | |
| Myasthenia gravis | 2/33 (6%) | |
| Cerebral venous sinus thrombosis | 1/33 (3%) | Five years prior to first included MRI |
| Transient ischaemic attack | 1/33 (3%) | Seven years prior to first included MRI |
| Pituitary adenoma | 1/33 (3%) | Treatment 11 years prior to first included MRI |
| **Treatment during stable phase** | | |
| Attack-preventing treatment | 30/33 (91%) | |
| Rituximab | 20/33 (61%) | |
| Azathioprine | 7/33 (21%) | |
| Prednisolone | 5/33 (15%) | |
| Mycophenolic acid | 4/33 (12%) | |
| Tocilizumab | 1/33 (3%) | |
| Glatiramer acetate | 1/33 (3%) | |
| Pain medication | 13/33 (39%) | |
| Gabapentin | 5/33 (15%) | |
| Oxycodone | 1/33 (3%) | |
| Metamizole | 1/33 (3%) | |
| Tilidine | 1/33 (3%) | |
| Ibuprofen | 1/33 (3%) | |
| Other | 4/33 (12%) | |
| Treatment for spasticity | 6/33 (18%) | |
| Baclofen | 3/33 (9%) | |
| Pramipexole | 2/33 (6%) | |
| Tizanidine | 1/33 (3%) | |
| Hyoscine butyl bromide | 1/33 (3%) | |
| Anxiolytic/antidepressant medication | 7/33 (21%) | |
| Pregabalin | 5/33 (15%) | |
| Mirtazapine | 2/33 (6%) | |
| Amitriptyline | 2/33 (6%) | |
| Treatment for hypertension | 10/33 (30%) | |
| **Reasons for study exclusion** | | |
| MOG-IgG-seropositive | 20 | |
| Double-seropositive (AQP4-IgG/MOG-IgG) | 1 | |
| AQP4-IgG-seronegative | 3 | |
| Recent relapses (< 1 year before MRI) | 3 | |
| Significant CNS comorbidity | 2 | Ischaemic stroke, suspected secondary MS |
| Unable to confirm stable disease course | 12 | |
| Unable to confirm AQP4-IgG serostatus | 5 | |

**Supplementary Table 2 | Brain Lesion Subsegmentation Values at Patient Baseline MRI**

| **Brain Lesion Location** | **Median Lesion Count [range]** | **Proportion of Patients with Lesions** | | **Lesion Volume (mL)** | |
|---|---|---|---|---|---|
| | | *n/N [decimal fraction]* | *95% CI* | *Mean* | *SD* |
| Non-specific WM | 4 [0 - 48] | 27/33 [0.82] | 0.69 - 0.95 | 0.469 | 0.768 |
| Juxtacortical | 1 [0 - 13] | 18/33 [54.5] | 0.38 - 0.72 | 0.127 | 0.245 |
| Lateral Ventricle | 3 [0 – 21] | 29/33 [87.9] | 0.51 - 0.83 | 1.244 | 2.094 |
| Diencephalic including 3rd Ventricle | 0 [0 – 3] | 6/33 [18.2] | 0.05 - 0.31 | 0.013 | 0.035 |
| Corpus Callosum | 0 [0 – 0] | 0/33 [0.0] | NA | 0.000 | 0.000 |
| Brainstem including 4th Ventricle | 0 [0 – 5] | 6/33 [18.2] | 0.05 - 0.31 | 0.024 | 0.087 |
| Cerebellum | 0 [0 – 1] | 1/33 [0.03] | -0.03 - 0.09 | 0.002 | 0.009 |
| CST Extensive | 0 [0 – 1] | 1/33 [0.03] | -0.03 - 0.09 | 0.002 | 0.009 |

**Supplementary Table 3 | Mean damage of cohort white matter tract regions extracted from longitudinal DWI in each individual patient**

| White Matter Tract Region | Mean Damage (%) [±SD, range] | Thresholded Mean Damage (%) [±SD, range] |
|---|---|---|
| Left CC | 6.8 [±4.9, 0.0 – 16.3] | 0.0 [±0.0, 0.0 – 0.0] |
| Right CC | 7.9 [±4.9, 0.0 – 21.7] | 0.0 [±0.0, 0.0 – 0.0] |
| Left Anterior Cingulum | 7.9 [±4.2, 0.0 – 23.5] | 0.3 [±2.0, 0.0 – 18.6] |
| Left Posterior Cingulum | 7.8 [±4.4, 0.9 – 18.9] | 0.4 [±1.3, 0.0 – 8.0] |
| Right Anterior Cingulum | 8.1 [±4.0, 0.2 – 19.9] | 0.2 [±1.2, 0.0 – 9.7] |
| Right Posterior Cingulum | 8.4 [±4.4, 1.6 – 19.5] | 0.1 [±0.8, 0 – 6.1] |
| **Left CST Brainstem Region** | **14.1 [±6.9, 0.0 – 37.5]** | **6.9 [±6.9, 0.0 – 36.6]** |
| Left CST Cortical Region | 9.6 [±5.3, 0.1 – 28.1] | 1.8 [±3.4, 0.0 – 14.2] |
| **Left CST Mid-Region** | **11.7 [±8.7, 0.0 – 62.0]** | **5.5 [±8.1, 0.0 – 58.7]** |
| Right CST Brainstem Region | 9.7 [±4.9, 0.2 – 25.3] | 1.8 [±3.7, 0.0 – 20.0] |
| Right CST Cortical Region | 8.6 [±5.3, 0.9 – 29.9] | 1.7 [±3.8, 0.0 – 20.8] |
| **Right CST Mid-Region** | **13.0 [±10.6, 0.6 – 51.3]** | **7.2 [±10.0, 0.0 – 47.2]** |
| Left Anterior IFOF | 7.4 [±3.0, 0.7 – 14.9] | 0.1 [±0.5, 0.0 – 3.6] |
| Left IFOF Mid-Region | 7.2 [±4.7, 0.0 – 21.1] | 0.5 [±1.5, 0.0 – 8.2] |
| Left Posterior IFOF | 7.7 [±3.6, 0.8 – 15.0] | 0.0 [±0.0, 0.0 – 0.0] |
| Right Anterior IFOF | 7.7 [±3.4, 1.9 – 22.4] | 0.3 [±1.2, 0.0 – 8.3] |
| Right IFOF Mid-Region | 7.3 [±5.1, 0.0 – 25.6] | 0.5 [±1.8, 0.0 – 11.3] |
| Right Posterior IFOF | 7.3 [±4.5, 0.1 – 24.3] | 0.2 [±1.0, 0.0 – 6.1] |
| Left OR Cortical Region | 7.7 [±4.6, 0.2 – 25.2] | 0.6 [±2.3, 0.0 – 18.3] |
| Left OR Mid-Region | 8.3 [±6.1, 0.0 – 26.5] | 1.6 [±2.7, 0.0 – 11.0] |
| Left OR Thalamic Region | 6.9 [±6.4, 0.0 – 25.7] | 1.1 [±2.6, 0.0 – 11.2] |
| Right OR Cortical Region | 7.7 [±4.4, 0.4 – 18.2] | 0.3 [±1.3, 0.0 – 7.4] |
| **Right OR Mid-Region** | **8.6 [±7.0, 0.0 – 30.9]** | **2.2 [±4.4, 0.0 – 22.6]** |
| Right OR Thalamic Region | 6.6 [±4.6, 0.0 – 18.2] | 0.3 [±1.2, 0.0 – 8.0] |
| Left Anterior TPOL | 7.7 [±5.8, 0.0 – 22.8] | 0.5 [±1.7, 0.0 – 8.4] |
| Left TPOL Mid-Region | 8.5 [±5.3, 0.0 – 21.3] | 0.8 [±2.1, 0.0 – 10.6] |
| Left Posterior TPOL | 7.6 [±4.1, 0.2 – 27.2] | 0.3 [±2.1, 0.0 – 19.0] |
| Right Anterior TPOL | 6.5 [±4.4, 0.0 – 18.4] | 0.1 [±0.6, 0.0 – 4.2] |
| Right TPOL Mid-Region | 7.9 [±6.6, 0.0 – 25.3] | 1.5 [±3.0, 0.0 – 14.3] |
| Right Posterior TPOL | 7.7 [±4.3, 0.9 – 19.1] | 0.4 [±1.2, 0.0 – 5.5] |
| Left Anterior TPFC | 7.8 [±3.4, 1.1 – 15.6] | 0.0 [±0.4, 0.0 – 3.8] |
| Left TPFC Mid-Region | 7.3 [±4.9, 0.0 – 24.7] | 0.2 [±1.5, 0.0 – 13.4] |
| **Left Posterior TPFC** | **10.6 [±9.4, 0.0 – 57.2]** | **3.4 [±7.6, 0.0 – 52.7]** |
| Right Anterior TPFC | 9.2 [±4.7, 0.2 – 20.8] | 0.1 [±0.8, 0.0 – 4.3] |
| Right TPFC Mid-Region | 8.3 [±5.2, 0.5 – 25.6] | 0.5 [±1.6, 0.0 – 7.7] |
| **Right Posterior TPFC** | **15.2 [±11.2, 0.0 – 59.4]** | **8.9 [±11.0, 0.0 – 56.0]** |

**Supplementary Table 4 | Cross-sectional analysis of first available DWI regional mean FA values in patients with the upper tertile of thalamic volumes versus the rest of the cohort**

| WM Fibre Bundle Region | Group 1 | Group 2 | Effect Size (Cohen's d) | Effect Size 95% CI | | Effect Size Magnitude |
|---|---|---|---|---|---|---|
| Left Anterior Cingulum | upper tertile | rest of cohort | 0.98 | 0.21 | 2 | large |
| Left Anterior IFOF | upper tertile | rest of cohort | 0.45 | -0.27 | 1.46 | small |
| Left Anterior TPFC | upper tertile | rest of cohort | 0.58 | -0.2 | 1.59 | moderate |
| Left Anterior TPOL | upper tertile | rest of cohort | 0.13 | -0.64 | 0.97 | negligible |
| Left CC | upper tertile | rest of cohort | 0.37 | -0.45 | 1.09 | small |
| Left CST Brainstem Region | upper tertile | rest of cohort | -0.03 | -1.05 | 0.81 | negligible |
| Left CST Cortical Region | upper tertile | rest of cohort | 0.32 | -0.47 | 1.74 | small |
| Left CST Mid-Region | upper tertile | rest of cohort | 0.09 | -0.68 | 0.83 | negligible |
| Left IFOF Mid-Region | upper tertile | rest of cohort | 0.58 | -0.05 | 1.38 | moderate |
| Left OR Cortical Region | upper tertile | rest of cohort | 0.59 | -0.11 | 1.47 | moderate |
| Left OR Mid-Region | upper tertile | rest of cohort | 0.68 | -0.01 | 1.66 | moderate |
| Left OR Thalamic Region | upper tertile | rest of cohort | 0.18 | -0.61 | 1.14 | negligible |
| Left Posterior Cingulum | upper tertile | rest of cohort | 1.16 | 0.4 | 2.16 | large |
| Left Posterior IFOF | upper tertile | rest of cohort | 0.75 | 0.08 | 1.49 | moderate |
| Left Posterior TPFC | upper tertile | rest of cohort | 0.27 | -0.42 | 1.05 | small |
| Left Posterior TPOL | upper tertile | rest of cohort | 0.65 | -0.08 | 1.58 | moderate |
| Left TPFC Mid-Region | upper tertile | rest of cohort | 0.19 | -0.64 | 0.99 | negligible |
| Left TPOL Mid-Region | upper tertile | rest of cohort | 0.83 | 0.09 | 1.72 | large |
| Right Anterior Cingulum | upper tertile | rest of cohort | 0.58 | -0.16 | 1.7 | moderate |
| Right Anterior IFOF | upper tertile | rest of cohort | 0.82 | 0.06 | 1.85 | large |
| Right Anterior TPFC | upper tertile | rest of cohort | 0.95 | 0.14 | 2.61 | large |

| | | | | | | |
|---|---|---|---|---|---|---|
| Right Anterior TPOL | upper tertile | rest of cohort | 0.18 | -0.59 | 1.1 | negligible |
| Right CC | upper tertile | rest of cohort | 0.62 | -0.12 | 1.32 | moderate |
| Right CST Brainstem Region | upper tertile | rest of cohort | 0.58 | -0.08 | 1.34 | moderate |
| Right CST Cortical Region | upper tertile | rest of cohort | 0.55 | -0.25 | 1.57 | moderate |
| Right CST Mid-Region | upper tertile | rest of cohort | -0.04 | -0.82 | 0.59 | negligible |
| Right IFOF Mid-Region | upper tertile | rest of cohort | 0.07 | -0.73 | 0.76 | negligible |
| Right OR Cortical Region | upper tertile | rest of cohort | 0.37 | -0.32 | 1.16 | small |
| Right OR Mid-Region | upper tertile | rest of cohort | 0.68 | -7.9e-4 | 1.52 | moderate |
| Right OR Thalamic Region | upper tertile | rest of cohort | 0.21 | -0.56 | 1.12 | small |
| Right Posterior Cingulum | upper tertile | rest of cohort | 1.33 | 0.54 | 2.52 | large |
| Right Posterior IFOF | upper tertile | rest of cohort | 0.68 | -0.07 | 1.96 | moderate |
| Right Posterior TPFC | upper tertile | rest of cohort | 0.88 | 0.08 | 2.02 | large |
| Right Posterior TPOL | upper tertile | rest of cohort | 0.39 | -0.29 | 1.32 | small |
| Right TPFC Mid-Region | upper tertile | rest of cohort | 0.16 | -0.59 | 1 | negligible |
| Right TPOL Mid-Region | upper tertile | rest of cohort | 0.63 | -0.08 | 1.53 | moderate |

**Supplementary Table 5 | Representative patients with highest thalamic volumes and the longitudinal change in their white matter fibre-bundle regions**

| Subject | Session | White Matter Tract Region | Mean FA | Delta Mean FA (Visit 2 – Visit 1) | Percent Change in FA from Visit 1 |
|---|---|---|---|---|---|
| 1 | 1 | Left CC | 0.73 | NA | NA |
| 1 | 1 | Right CC | 0.69 | NA | NA |
| 1 | 1 | Left Anterior Cingulum | 0.57 | NA | NA |
| 1 | 1 | Left Posterior Cingulum | 0.51 | NA | NA |
| 1 | 1 | Right Anterior Cingulum | 0.52 | NA | NA |
| 1 | 1 | Right Posterior Cingulum | 0.55 | NA | NA |
| 1 | 1 | Left CST Brainstem Region | 0.4 | NA | NA |
| 1 | 1 | Left CST Cortical Region | 0.43 | NA | NA |
| 1 | 1 | Left CST Mid-Region | 0.66 | NA | NA |
| 1 | 1 | Right CST Brainstem Region | 0.45 | NA | NA |
| 1 | 1 | Right CST Cortical Region | 0.45 | NA | NA |
| 1 | 1 | Right CST Mid-Region | 0.72 | NA | NA |
| 1 | 1 | Left Anterior IFOF | 0.41 | NA | NA |
| 1 | 1 | Left IFOF Mid-Region | 0.46 | NA | NA |
| 1 | 1 | Left Posterior IFOF | 0.46 | NA | NA |
| 1 | 1 | Right Anterior IFOF | 0.42 | NA | NA |
| 1 | 1 | Right IFOF Mid-Region | 0.47 | NA | NA |
| 1 | 1 | Right Posterior IFOF | 0.49 | NA | NA |
| 1 | 1 | Left OR Cortical Region | 0.49 | NA | NA |
| 1 | 1 | Left OR Mid-Region | 0.55 | NA | NA |
| 1 | 1 | Left OR Thalamic Region | 0.54 | NA | NA |
| 1 | 1 | Right OR Cortical Region | 0.41 | NA | NA |
| 1 | 1 | Right OR Mid-Region | 0.53 | NA | NA |
| 1 | 1 | Right OR Thalamic Region | 0.5 | NA | NA |
| 1 | 1 | Left Anterior TPOL | 0.53 | NA | NA |
| 1 | 1 | Left TPOL Mid-Region | 0.55 | NA | NA |
| 1 | 1 | Left Posterior TPOL | 0.48 | NA | NA |
| 1 | 1 | Right Anterior TPOL | 0.5 | NA | NA |
| 1 | 1 | Right TPOL Mid-Region | 0.53 | NA | NA |
| 1 | 1 | Right Posterior TPOL | 0.43 | NA | NA |
| 1 | 1 | Left Anterior TPFC | 0.39 | NA | NA |
| 1 | 1 | Left TPFC Mid-Region | 0.6 | NA | NA |
| 1 | 1 | Left Posterior TPFC | 0.41 | NA | NA |
| 1 | 1 | Right Anterior TPFC | 0.38 | NA | NA |
| 1 | 1 | Right TPFC Mid-Region | 0.56 | NA | NA |

| | | | | | |
|---|---|---|---|---|---|
| 1 | 1 | Right Posterior TPFC | 0.3 | NA | NA |
| 1 | 2 | Left CC | 0.71 | -0.02 | -2.74 |
| 1 | 2 | Right CC | 0.68 | -0.01 | -1.44 |
| 1 | 2 | Left Anterior Cingulum | 0.52 | -0.04 | -7.08 |
| 1 | 2 | Left Posterior Cingulum | 0.47 | -0.04 | -7.8 |
| 1 | 2 | Right Anterior Cingulum | 0.47 | -0.05 | -9.65 |
| 1 | 2 | Right Posterior Cingulum | 0.49 | -0.05 | -9.12 |
| 1 | 2 | **Left CST Brainstem Region** | **0.41** | **0.01** | **2.47** |
| 1 | 2 | Left CST Cortical Region | 0.41 | -0.02 | -4.64 |
| 1 | 2 | Left CST Mid-Region | 0.64 | -0.02 | -3.03 |
| 1 | 2 | Right CST Brainstem Region | 0.43 | -0.02 | -4.41 |
| 1 | 2 | Right CST Cortical Region | 0.43 | -0.02 | -4.43 |
| 1 | 2 | Right CST Mid-Region | 0.68 | -0.04 | -5.59 |
| 1 | 2 | Left Anterior IFOF | 0.41 | 0 | 0 |
| 1 | 2 | **Left IFOF Mid-Region** | **0.48** | **0.01** | **2.15** |
| 1 | 2 | Left Posterior IFOF | 0.46 | 0 | 0 |
| 1 | 2 | Right Anterior IFOF | 0.42 | 0 | 0 |
| 1 | 2 | **Right IFOF Mid-Region** | **0.48** | **0.01** | **2.13** |
| 1 | 2 | Right Posterior IFOF | 0.47 | -0.02 | -4.09 |
| 1 | 2 | Left OR Cortical Region | 0.49 | 0 | 0 |
| 1 | 2 | **Left OR Mid-Region** | **0.57** | **0.01** | **1.81** |
| 1 | 2 | Left OR Thalamic Region | 0.54 | 0 | 0 |
| 1 | 2 | Right OR Cortical Region | 0.4 | -0.01 | -2.46 |
| 1 | 2 | Right OR Mid-Region | 0.52 | -0.01 | -1.88 |
| 1 | 2 | **Right OR Thalamic Region** | **0.51** | **0.01** | **1.99** |
| 1 | 2 | Left Anterior TPOL | 0.52 | 0 | 0 |
| 1 | 2 | **Left TPOL Mid-Region** | **0.57** | **0.01** | **1.81** |
| 1 | 2 | Left Posterior TPOL | 0.48 | 0 | 0 |
| 1 | 2 | **Right Anterior TPOL** | **0.51** | **0.01** | **1.98** |
| 1 | 2 | Right TPOL Mid-Region | 0.52 | -0.01 | -1.87 |
| 1 | 2 | Right Posterior TPOL | 0.43 | -0.01 | -2.3 |
| 1 | 2 | Left Anterior TPFC | 0.38 | -0.01 | -2.57 |
| 1 | 2 | Left TPFC Mid-Region | 0.6 | 0 | 0 |
| 1 | 2 | Left Posterior TPFC | 0.34 | -0.08 | -19.35 |
| 1 | 2 | Right Anterior TPFC | 0.35 | -0.03 | -7.87 |
| 1 | 2 | Right TPFC Mid-Region | 0.55 | -0.01 | -1.79 |
| 1 | 2 | Right Posterior TPFC | 0.22 | -0.07 | -23.43 |
| 2 | 1 | Left CC | 0.71 | NA | NA |
| 2 | 1 | Right CC | 0.67 | NA | NA |
| 2 | 1 | Left Anterior Cingulum | 0.59 | NA | NA |

| | | | | | |
|---|---|---|---|---|---|
| 2 | 1 | Left Posterior Cingulum | 0.51 | NA | NA |
| 2 | 1 | Right Anterior Cingulum | 0.52 | NA | NA |
| 2 | 1 | Right Posterior Cingulum | 0.57 | NA | NA |
| 2 | 1 | Left CST Brainstem Region | 0.42 | NA | NA |
| 2 | 1 | Left CST Cortical Region | 0.37 | NA | NA |
| 2 | 1 | Left CST Mid-Region | 0.67 | NA | NA |
| 2 | 1 | Right CST Brainstem Region | 0.45 | NA | NA |
| 2 | 1 | Right CST Cortical Region | 0.45 | NA | NA |
| 2 | 1 | Right CST Mid-Region | 0.69 | NA | NA |
| 2 | 1 | Left Anterior IFOF | 0.45 | NA | NA |
| 2 | 1 | Left IFOF Mid-Region | 0.51 | NA | NA |
| 2 | 1 | Left Posterior IFOF | 0.41 | NA | NA |
| 2 | 1 | Right Anterior IFOF | 0.45 | NA | NA |
| 2 | 1 | Right IFOF Mid-Region | 0.47 | NA | NA |
| 2 | 1 | Right Posterior IFOF | 0.45 | NA | NA |
| 2 | 1 | Left OR Cortical Region | 0.45 | NA | NA |
| 2 | 1 | Left OR Mid-Region | 0.52 | NA | NA |
| 2 | 1 | Left OR Thalamic Region | 0.54 | NA | NA |
| 2 | 1 | Right OR Cortical Region | 0.41 | NA | NA |
| 2 | 1 | Right OR Mid-Region | 0.54 | NA | NA |
| 2 | 1 | Right OR Thalamic Region | 0.51 | NA | NA |
| 2 | 1 | Left Anterior TPOL | 0.52 | NA | NA |
| 2 | 1 | Left TPOL Mid-Region | 0.52 | NA | NA |
| 2 | 1 | Left Posterior TPOL | 0.44 | NA | NA |
| 2 | 1 | Right Anterior TPOL | 0.51 | NA | NA |
| 2 | 1 | Right TPOL Mid-Region | 0.54 | NA | NA |
| 2 | 1 | Right Posterior TPOL | 0.43 | NA | NA |
| 2 | 1 | Left Anterior TPFC | 0.39 | NA | NA |
| 2 | 1 | Left TPFC Mid-Region | 0.58 | NA | NA |
| 2 | 1 | Left Posterior TPFC | 0.4 | NA | NA |
| 2 | 1 | Right Anterior TPFC | 0.36 | NA | NA |
| 2 | 1 | Right TPFC Mid-Region | 0.57 | NA | NA |
| 2 | 1 | Right Posterior TPFC | 0.29 | NA | NA |
| 2 | 2 | Left CC | 0.7 | 0 | 0 |
| 2 | 2 | **Right CC** | **0.69** | **0.01** | **1.48** |
| 2 | 2 | Left Anterior Cingulum | 0.57 | -0.02 | -3.4 |
| 2 | 2 | Left Posterior Cingulum | 0.51 | 0 | 0 |
| 2 | 2 | **Right Anterior Cingulum** | **0.52** | **0.01** | **1.93** |
| 2 | 2 | **Right Posterior Cingulum** | **0.58** | **0.01** | **1.76** |
| 2 | 2 | Left CST Brainstem Region | 0.41 | 0 | 0 |

| | | | | | |
|---|---|---|---|---|---|
| 2 | 2 | Left CST Cortical Region | 0.37 | 0 | 0 |
| 2 | 2 | Left CST Mid-Region | 0.66 | -0.01 | -1.49 |
| 2 | 2 | Right CST Brainstem Region | 0.44 | -0.01 | -2.21 |
| **2** | **2** | **Right CST Cortical Region** | **0.46** | **0.01** | **2.23** |
| 2 | 2 | **Right CST Mid-Region** | **0.7** | **0.01** | **1.45** |
| 2 | 2 | Left Anterior IFOF | 0.43 | -0.01 | -2.24 |
| 2 | 2 | Left IFOF Mid-Region | 0.5 | -0.01 | -1.96 |
| 2 | 2 | Left Posterior IFOF | 0.4 | -0.01 | -2.42 |
| 2 | 2 | Right Anterior IFOF | 0.44 | 0 | 0 |
| 2 | 2 | **Right IFOF Mid-Region** | **0.49** | **0.02** | **4.27** |
| 2 | 2 | **Right Posterior IFOF** | **0.45** | **0.01** | **2.24** |
| 2 | 2 | Left OR Cortical Region | 0.43 | -0.02 | -4.45 |
| 2 | 2 | Left OR Mid-Region | 0.5 | -0.01 | -1.93 |
| 2 | 2 | Left OR Thalamic Region | 0.53 | -0.01 | -1.86 |
| 2 | 2 | Right OR Cortical Region | 0.41 | 0 | 0 |
| 2 | 2 | Right OR Mid-Region | 0.54 | -0.01 | -1.84 |
| 2 | 2 | **Right OR Thalamic Region** | **0.52** | **0.01** | **1.96** |
| 2 | 2 | Left Anterior TPOL | 0.51 | -0.01 | -1.94 |
| 2 | 2 | Left TPOL Mid-Region | 0.5 | -0.02 | -3.87 |
| 2 | 2 | Left Posterior TPOL | 0.42 | -0.02 | -4.56 |
| 2 | 2 | **Right Anterior TPOL** | **0.52** | **0.01** | **1.97** |
| 2 | 2 | Right TPOL Mid-Region | 0.53 | -0.01 | -1.86 |
| 2 | 2 | Right Posterior TPOL | 0.43 | 0 | 0 |
| 2 | 2 | Left Anterior TPFC | 0.39 | -0.01 | -2.54 |
| 2 | 2 | Left TPFC Mid-Region | 0.58 | -0.01 | -1.71 |
| 2 | 2 | Left Posterior TPFC | 0.38 | -0.01 | -2.51 |
| 2 | 2 | Right Anterior TPFC | 0.35 | 0 | 0 |
| 2 | 2 | Right TPFC Mid-Region | 0.57 | 0 | 0 |
| 2 | 2 | Right Posterior TPFC | 0.29 | 0 | 0 |
